# Hematologic dynamics during pregnancy and their association with obstetric complications: a retrospective cohort study

**DOI:** 10.1101/2025.02.13.25322250

**Authors:** Veronica Tozzo, Rachel Petherbridge, Kaitlyn James, Sarah Hsu, Chloe Michalopoulos, Brody H. Foy, Tanayott Thaweethai, Christopher Mow, Jacqueline Maya, Carolina Batlle Camero, Lydia Shook, Kathryn J. Gray, Logan Mauney, John M. Higgins, Camille E. Powe

## Abstract

**Objectives:** Pregnancy alters hematologic state as measured by complete blood counts (CBC), but the longitudinal changes in CBC indices that define healthy pregnancies are not well established. Our objectives were (1) to define gestational age-specific reference intervals for CBCs and their longitudinal changes in a large United States-based cohort and (2) to use these reference intervals to examine associations between extreme CBC values and changes and risk of obstetric complications.

**Design:** Retrospective cohort study including electronic health record-based discovery and validation cohorts.

**Setting:** Academic medical center and affiliated health system in the United States between 1998 and 2022.

**Participants:** Individuals with singleton pregnancies delivering after 30 weeks’ gestation who presented for prenatal care prior to 20 weeks’. There were 45,992 pregnancies in the discovery cohort, 18% of whom had complications, and 50,603 in the validation cohort, 22% with complications.

**Main outcome measures:** Composite outcome (hypertensive disorder of pregnancy, small for gestational age birthweight or preterm birth) and its individual components. We analyzed associations between CBC results and outcomes using generalized estimating equations for logistic regression with Bonferroni correction for multiple hypothesis testing.

**Results:** Hematocrit, hemoglobin, and red cell count values above their reference intervals were associated with increased risk of the composite obstetric complication: OR [95% CI] of 1.4 [1.2, 1.6] p=1.8*x*10^−5^ for hematocrit; 1.7[1.4, 1.9] p=1.4*x*10^−10^ for hemoglobin; and 1.6[1.4, 1.9] p=3.9×10^−9^ for red cell count. Extreme increase in hemoglobin (>0.67 g/dL) or red cell count (>0.07 10^6^/mm^3^) between 7-14 weeks’ and 26-29 weeks’ gestation was associated with increased risk for preterm birth (OR [95% CI] for hemoglobin 2.0[1.6, 2.6] p=2*x*10^−8^ and red cell count: 2.1[1.7, 2.6] p=9*x*10^−14^). Reference intervals in this cohort were often wider than those previously published for mean red cell volume, mean red cell hemoglobin, red cell count, and mean red cell hemoglobin concentration.

**Conclusions:** Elevated measures of red blood cell count and large intra-pregnancy increases in those measures are associated with subsequent obstetric complications.

**Summary box:** *What is already known on this topic:* Pregnancy causes significant changes in hematologic state as measured by routine complete blood counts (CBCs).

*What this study adds:* We studied more than 95,000 pregnancies and found that elevations (both absolute and relative to baseline 1^st^ trimester values) in hematocrit, hemoglobin, and red cell count are associated with adverse pregnancy outcomes including hypertensive disorder of pregnancy, small for gestational age birthweight, and preterm birth.

*Meaning:* Routinely collected prenatal CBCs may provide an opportunity to identify those at risk for obstetric complications.

## Introduction

Pregnancy leads to alterations in the hematologic system^1,2^ that are reflected in changes to complete blood counts (CBCs) routinely collected during prenatal care^3,4^, but consensus is lacking on what constitutes normal physiological change in pregnancy^5,6^. Abnormal CBC values or longitudinal changes have been shown outside pregnancy to be useful for clinical risk stratification^7^, but they have not been systematically evaluated as risk markers^8–10^ in pregnancy. With ∼85% of pregnant individuals in the United States (US) receiving longitudinal prenatal care that includes repeated CBC assessments^11^, there is a potential missed opportunity for early identification of obstetric complications.

Pregnancy-specific reference intervals are available but are derived from cross-sectional studies^12–17^ and meta-analyses of studies with small sample sizes^3,4,16,18,19^. Utilization of pregnancy-specific reference ranges by clinicians is limited, and their clinical utility has not been definitively demonstrated^3,4,6^ with the exception in some cases of pregnancy-specific lower limits of hemoglobin used to diagnose anemia^20^. Intra-pregnancy changes in CBCs have not been systematically investigated, with analysis limited to small studies^15,18,19,21,22^ or cross-sectional electronic health record (EHR) analysis^5,17,23^, and there has been no evaluation of intra-pregnancy CBC changes for routine risk stratification of pregnancy.

Here, we derive gestational age-specific reference intervals for CBC indices and their intra-pregnancy longitudinal changes in a large US-based cohort. We then use these reference intervals to systematically evaluate the associations of extreme results and rare dynamics with pregnancy complications.

## Methods

Study methods are summarized below; additional details appear in the **Supplementary Methods**.

### Study setting and cohort description

Our discovery cohort was the EHR-based Maternal Health Cohort which includes all pregnant patients who received prenatal care or had a delivery at Massachusetts General Hospital between 1998-2016^24–26^. We developed an out-of-sample validation cohort of pregnancies delivered across the Mass General Brigham (MGB) health system between 2016-2022. The MGB Institutional Review Board approved the study and waived the requirement for informed consent. For this analysis, we included singleton pregnancies ending in livebirth with a first prenatal visit before 20 weeks’ gestation and with data on at least one CBC checked in pregnancy. We excluded those who had known blood-related disorders (see Supplementary Methods for details), transfusions during or within 8 weeks prior to pregnancy, who delivered prior to 30 weeks’ gestation, or who were missing data on birth weight or neonatal sex (required for determining the primary outcome).

Pregnancies meeting inclusion criteria were divided into those with and without the following complications: preterm birth (<37 weeks’ gestation), hypertensive disorders of pregnancy (HDP) (including both gestational hypertension and preeclampsia, based on outpatient blood pressures meeting ACOG criteria^27^ and/or diagnosis documented in the delivery record), small for gestational age birth weight (SGA) (birth weight <10^th^ percentile for gestational age and sex^28^), and transfusion at delivery or within 8 weeks postpartum. We also examined preeclampsia, ascertained among HDP pregnancies based on laboratory criteria.^27^ We defined a composite primary outcome including HDP (inclusive of preeclampsia), preterm birth, or SGA and also examined individual outcomes. We used similar exclusion criteria and adverse outcome definitions in the validation cohort, except that HDP status was ascertained based on labor and delivery encounter ICD-10 codes^29^. We considered hematocrit (HCT), hemoglobin (HGB), white blood cell count (WBC), red cell count (RBC), platelet count (PLT), mean red cell volume (MCV), mean red cell hemoglobin content (MCH), red cell distribution width (RDW), and mean red cell hemoglobin concentration (MCHC). Based on the timing of routine CBC screening (**Figure S1**), we considered three pregnancy timepoints: 7-14 weeks’ gestation, 26-29 weeks’ gestation, and 0 to 7 days prior to term delivery (≥37 weeks’ gestation). We also considered pre-pregnancy CBCs for individuals who had ≤2 CBCs at least 6 months apart in the previous five years with no ICD codes indicating an illness or pregnancy at the time of the CBC.

### Statistical analysis

We defined gestational age-specific reference intervals for pregnancies without complications in the discovery cohort as the 2.5th and 97.5th percentiles of each CBC index at each timepoint. Literature reference intervals were collected from two commonly used online sources^30,31^ derived from meta-analyses^4^ (**Table S1)**. In sensitivity analyses, we determined gestational age-specific intervals in pregnancies without anemia (defined with ACOG criteria^20,32^) and in a subset of term pregnancies without prior chronic conditions or pregnancy-related issues recorded in the EHR.

We also investigated intra-pregnancy longitudinal changes in the discovery cohort by analyzing a subset of term pregnancies without complications for which CBCs were available at all 4 examined time points (pre-pregnancy, 7-14 weeks’ gestation, 26-29 weeks’ gestation, and pre-delivery). Intra-pregnancy CBC changes were analyzed by calculating simple differences between successive CBC measurements. Also, to focus on intra-pregnancy changes rather than differences in visit timing, we fit linear mixed-effects models adjusted for age and 1^st^ trimester BMI^26^, adding random effects for individual pregnancy, individual participant, parity, delivery year, and prenatal care site. These models enabled projection of CBC values to the same gestational age for all subjects. See **Supplementary Methods** for more detail on the projection modeling. Note that the projection models were used only for exploration, and all analysis of associations between longitudinal CBC changes and pregnancy outcomes used unadjusted raw CBC measurements in both the discovery and validation cohorts. In sensitivity analysis, we expanded to include all pregnancies with at least two consecutive time points, allowing for some missing data. The magnitude of intra-pregnancy longitudinal changes was compared to the magnitude of biological variation expected in non-pregnant adults^33^.

Odds ratios (OR) for adverse outcomes were computed with generalized estimating equations (GEE) for logistic regression in order to account for lack of independence between pregnancies in the same individual^34–36^. ORs were adjusted for 1^st^ trimester BMI, age, insurance status, race/ethnicity, and parity. Because we evaluated the risk at 26-29 weeks’ gestation, we excluded pregnancies with delivery <30 weeks’ gestation or HDP where the first high blood pressure occurred at <30 weeks’ gestation. Exposures were defined as a CBC result falling outside the 26-29 weeks’ reference interval or a rare intra-pregnancy longitudinal change in a CBC index.

Rare longitudinal changes were defined in the discovery cohort as a change in a CBC index between 7-14 weeks’ and 26-29 weeks’ gestation that was extreme in terms of both magnitude and direction. Direction was chosen as the least frequent variation exceeding established non-pregnant biological variation. Thresholds for these were then determined by analyzing all percentiles of the distribution of changes observed between 7-14 and 26-29 weeks’ gestation in the discovery cohort, and then choosing the threshold with the highest positive predictive value (PPV) and significant OR for each outcome. We focused on PPV because it helps assess a marker’s potential clinical significance, which typically requires that the PPV significantly exceed the prevalence of the outcome being predicted. All thresholds with PPVs whose confidence intervals did not include the prevalence of a given outcome in the discovery cohort potentially provided novel prognostic value and were tested in the validation cohort.

Bonferroni correction of alpha = 0.05 was used to account for multiple hypothesis testing where appropriate, with the corresponding significance level specified in individual results. The analysis was performed in Python 3.11.3 and R version 4.3.1.

## Results

### Study population

The discovery cohort consisted of 45,992 pregnancies in 32,731 individuals (**Figure 1)**. There were 37,709 (82%) term pregnancies without relevant complications and 8,283 (18%) pregnancies with complications (**Table S2**). Pregnancies with and without complications had similar baseline characteristics (**Table 1**). In the validation cohort, 50,603 pregnancies met inclusion criteria (**Table S3**).

**Table 1.** Characteristics of the discovery cohort.

| Variable | Pregnancies without complications | Pregnancies with complications |
| --- | --- | --- |
| **Pregnancies N (%)** | 37709 (100) | 8283 (100) |
| **Age, years, Mean (SD)** | 32 (6) | 31 (6) |
| **Married or Partnered, N (%)** | 26938 (72) | 5664 (68) |
| **Insurance status, N (%)** |  |  |
| *Public* | 9400 (25) | 2232 (27) |
| *Private* | 24926 (66) | 5396 (65) |
| *None/limited* | 3383 (9) | 655 (8) |
| **Self-reported race, N(%)** |  |  |
| *Asian* | 3256 (9) | 829 (10) |
| *Black* | 2344 (6) | 657 (8) |
| *Latina* | 4659 (12) | 962 (12) |
| *White* | 22783 (60) | 4844 (58) |
| *None of the above* | 4667 (13) | 991 (12) |
| **Nulliparous, N (%)** | 16709 (44) | 4816 (58) |
| **Gestational week at delivery, weeks, Mean (SD)** | 40 (1) | 38 (2) |
| **Prenatal BMI at 12 weeks, kg/m<sup>2</sup>, mean (SD)** | 25.2 (5.0) | 25.8 (5.6) |
| **Time of CBC measurement** |  |  |
| *Pre – pregnancy* | 3980 (11) | 1057 (13) |
| *7-14 weeks' gestation* | 29126 (77) | 6365 (77) |
| *26-29 weeks' gestation* | 27941 (74) | 6218 (75) |
| *Pre-delivery* | 37528 (100) | 8260 (100) |
| *Pre-pregnancy & 7-14 weeks' gestation* | 3333 (9) | 868 (10) |
| *7-14 weeks & 26-29 weeks' gestation* | 24125 (64) | 5291 (64) |
| *26-29 weeks & Pre-delivery* | 27795 (74) | 6202 (74) |
| *All time points* | 2791 (7) | 729 (9) |

BMI at 12 weeks' gestation was either measured, interpolated from available measurements, or extrapolated based on a population model. When no BMI measurement was available, it was treated as missing. See **Supplementary Methods** for more details. CBC – Complete Blood Count. Complications –hypertensive disorder of pregnancy, preeclampsia, small for gestational age, preterm delivery at < 37 weeks' gestation, transfusion at or within 8 weeks after delivery.

**Figure 1.**
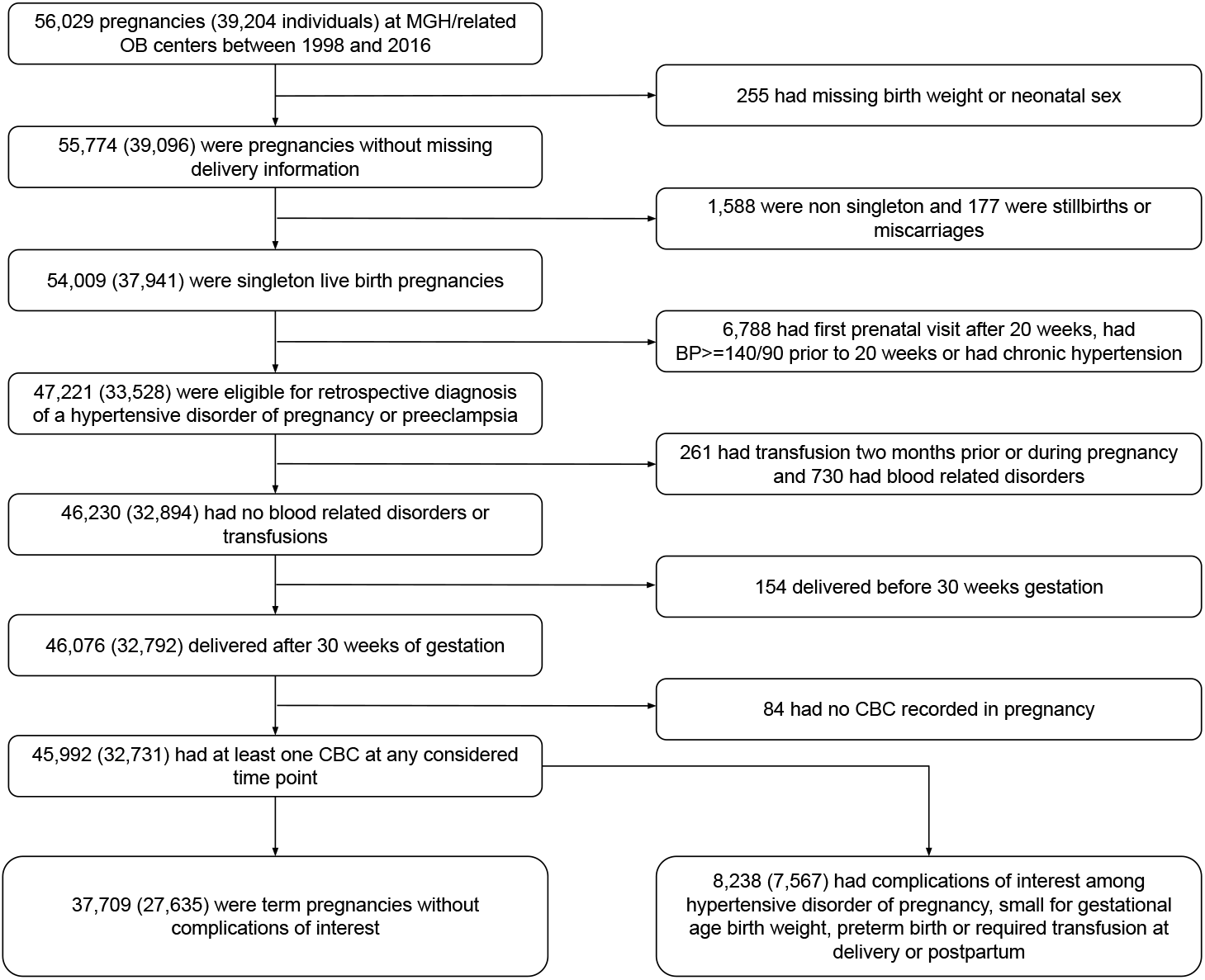
Selection of individuals from the Maternal Health Cohort.

### Multiple CBC reference intervals differ from those in commonly-cited literature

Reference intervals are defined by studying healthy populations, with inclusion and exclusion criteria determined by the specific diagnostic and demogaphic scenario^37^. Reference intervals for pregnancy should be based on distributions of test results obtained for pregnancies without complications. We analyzed these distributions in our discovery cohort (**Figure 1, Table 1**) and compared trimester-specific CBC index reference intervals to those in commonly-cited literature and to those in regular use for non-pregnant adult females (**Figure 2, Table S1**). RBC, MCV, MCH and MCHC reference intervals showed notable differences, with only 75, 74, 30, and 52% of uncomplicated pregnancies in our cohort falling within literature reference intervals throughout gestation, meaning that according to current practices at least 25% and as many as 70% of uncomplicated pregnancies had at least one abnormal result for each of these CBC indices. See Supplementary **Figures S2** and **S3** for more detail.

**Figure 2.**
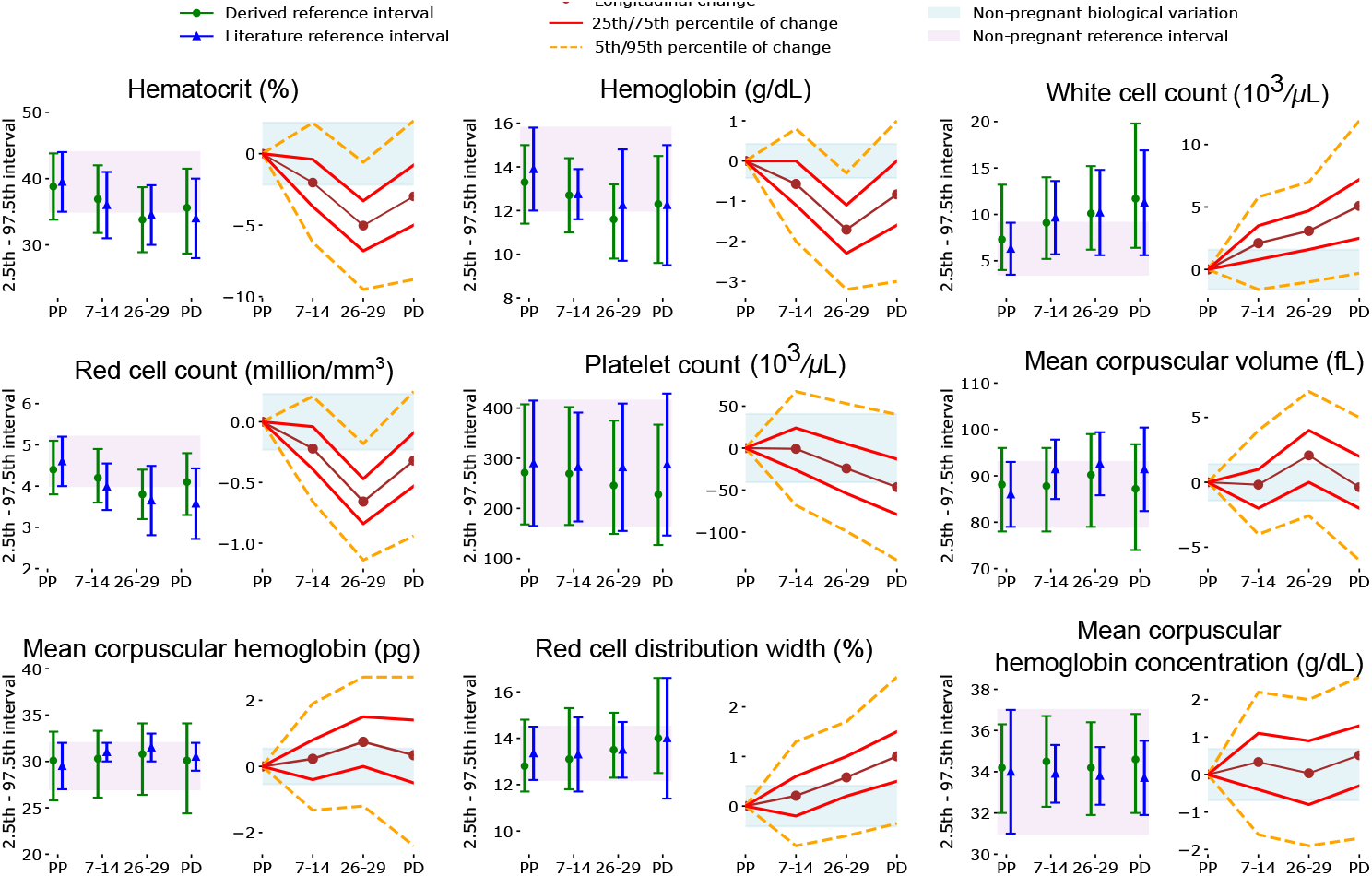
Reference intervals and average values for CBC indices vary substantially across gestation, with high inter-individual variability. For each marker, on the left are the gestational age-specific reference intervals from the Maternal Health Cohort (N=37,709, green error bars with round marker), in comparison with literature trimester-specific reference intervals (blue error bars with triangle). On the right are the intra-pregnancy longitudinal changes relative to pre-pregnancy baseline in N=2791 pregnancies for which CBCs were available at all considered time points. Literature and non-pregnant reference intervals were retrieved from prior publications^30,31^. PP is Pre-Pregnancy, and PD is Pre-Delivery at term.

### Elevated red cell indices are associated with complications

We investigated the use of the 26-29 weeks’ gestation reference intervals for the identification of pregnancies at elevated risk of subsequent adverse outcomes. Pregnancies with HGB, HCT, or RBC above the reference interval at 26-29 weeks’ gestation were significantly more likely to have the composite outcome (p<1.5*x*10^−4^, **Figure 3**): OR 1.4 [1.2, 1.7] p=1.8*x*10^−5^ for HCT, 1.7 [1.4, 1.9] p=1.4*x*10^−10^ for HGB, and 1.6 [1.4, 1.9] p=3.9*x*10^−9^ for RBC (**Table S7**). Elevated HGB and RBC at 26-29 weeks’ gestation were associated with HDP (1.7 [1.4, 2.2], p=7*x*10^−7^ HGB and 1.7 [1.4, 2.2] p=1.8*x*10^−6^ RBC), preeclampsia (1.9 [1.4, 2.5], p=1.2*x*10^−5^ HGB and 2.1 [1.6, 2.8] p=1*x*10^−7^ RBC), and preterm birth (1.8 [1.4, 2.3], p=8×10^−7^ HGB and 2.0 [1.6, 2.5] p=2.5*x*10^−9^ RBC) when these outcomes were examined individually (**Figure 3, Table S8-S12**). Values below the 26-29 weeks’ gestation reference interval were not significantly associated with the composite outcome or its individual components. As expected, peripartum transfusion was significantly associated with lower HGB (OR for HGB below the reference range: 3.4 [2.0, 5.8] p=4*x*10^−6^, **Figure 3, Table S10**). Results in the validation cohort were consistent with those in the primary analysis **(Table S13**).

**Figure 3.**
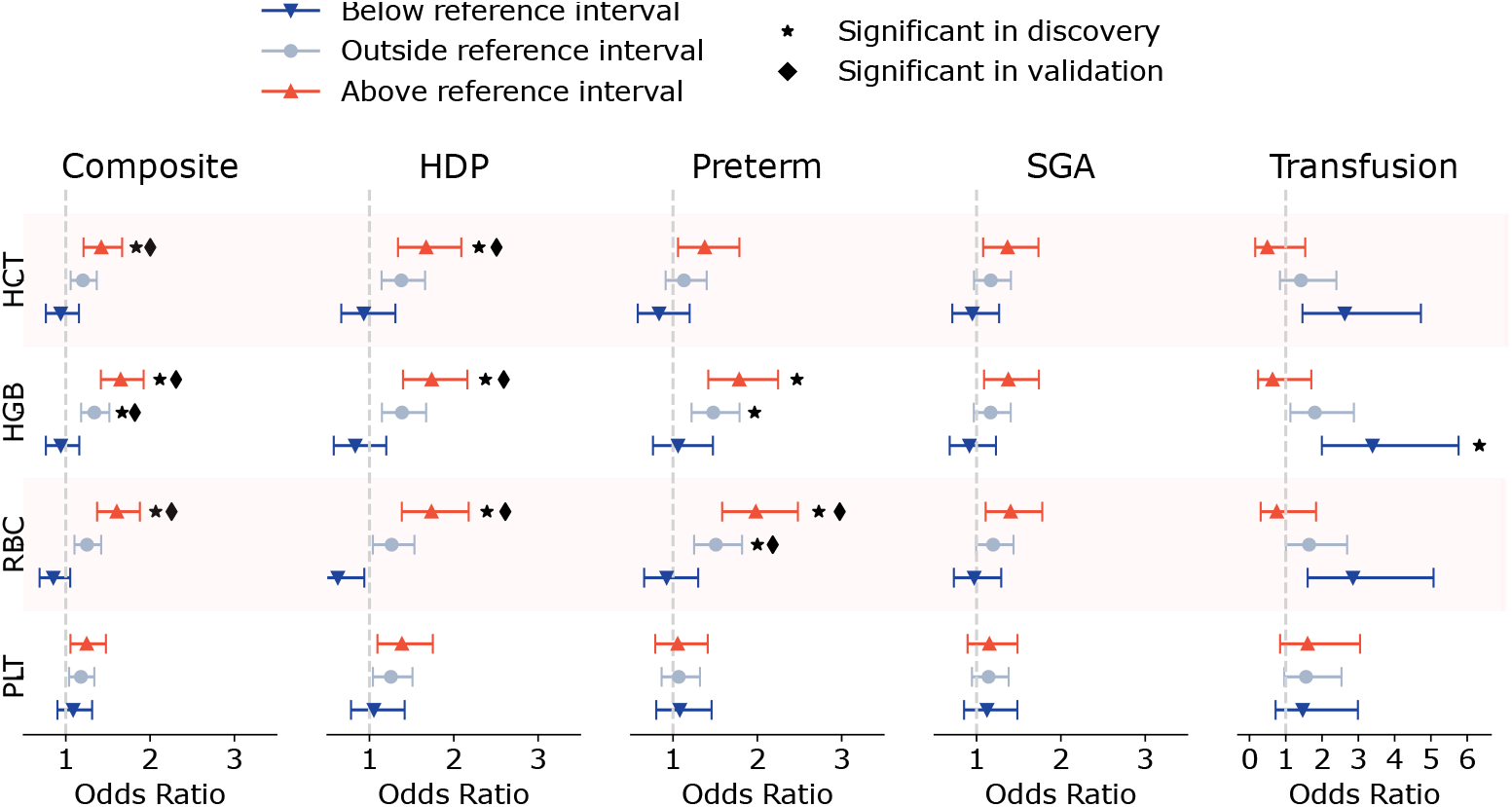
CBC values above their reference intervals are associated with complications. Odds ratios for developing the composite outcome (hypertensive disorders of pregnancy [HDP], small for gestational age birth weight [SGA], preterm birth) and individual complications are shown. Pregnancies with a CBC at 26-29 weeks’ gestation were considered (N=34,159), and those with preterm birth at <30 weeks’ gestation or evidence of HDP or preeclampsia before 29 weeks were excluded (N=297). Odds ratios are marked as significant in discovery or validation if the p-value is below 0.0002, reflecting Bonferroni correction.

### CBC indices change dramatically during uncomplicated pregnancies

In our analysis of the subset of the uncomplicated term pregnancies that had all CBCs available from the 4 examined timepoints (including prior to pregnancy and 3 pregnancy timepoints), CBC indices changed dramatically over time (**Figure 2** and **Table S4**), with 25 of 27 intra-pregnancy CBC index changes reaching statistical significance (p<0.002). The initial changes (pre-pregnancy to 7-14 weeks’ gestation) for PLT and MCV were the only exceptions. For all CBC indices, less than 1 in 3 and as few as 1 in 100 pregnancies changed during gestation by less than what is defined as healthy biological variation outside pregnancy (**Table S5**). Most CBC indices decreased initially between pre-pregnancy and the 1^st^ trimester. Then, between 7-14 weeks’ and 26-29 weeks’ gestation, HGB, RBC, and HCT decreased by an amount greater than biological variation in 82%, 81%, and 66% of pregnancies. WBC and PLT remained within biological variation in 54% and 66%, while MCV, MCH, RDW, and MCHC were typically either stable or increased during the same gestational time period. From 26-29 weeks’ gestation to pre-delivery (at term), HCT, HGB, WBC, RBC, RDW, and MCHC increased or were stable in ∼90% of pregnancies, while PLT, MCV, and MCH decreased or were stable (**Table S5**). Sensitivity analyses including pregnancies with missing CBC data from one or two timepoints were consistent with the primary analyses (**Table S6)**.

### Rare CBC index changes are associated with complications

Given the large variation in CBC indices during pregnancy, we explored in participants who had a CBC at 7-14 and 26-29 weeks’ gestation (N=29,416 discovery, N=50,603 validation) whether benchmarks for changes in CBC indices could also help identify pregnancies at risk of subsequent complications. We defined rare longitudinal changes for each CBC index based on the distribution of these changes in our study cohort. Changes less than biological variation were treated as stable. Between 7-14 and 26-29 weeks’ gestation, the least frequent changes were an increase in HGB (2%), HCT (1.3%), RBC (0.4%), PLT (4%), or MCHC (18%), and a decrease in WBC (8.4%), MCV (5.7%), MCH (13.4%), or RDW (10.1%) (**Figure 2** and **Table S5**). Rare changes were associated with increased risk of subsequent complications. Between 7-14 and 26-29 weeks’ gestation, an increase of >1.8% in HCT was associated with the composite outcome, as was an increase of >0.67 g/dL in HGB, an increase >0.07 10^6^/mm^3^ in RBC, a decrease of less than 0.05 10^3^/µl or an increase in PLT, and a decrease in MCV greater than 0.75 fL (**Table 2, Table S14**). No significant associations were found between rare longitudinal changes and peripartum transfusion (**Table S14**). Consistent results were found in the validation cohort (**Table S2 and S15-S18**). For the majority of pregnancies with complications identified by these rare changes, the relevant CBC index was within the 26-29 weeks’ gestation reference interval and was therefore unremarkable on its own (**Table 2**).

**Table 2.**
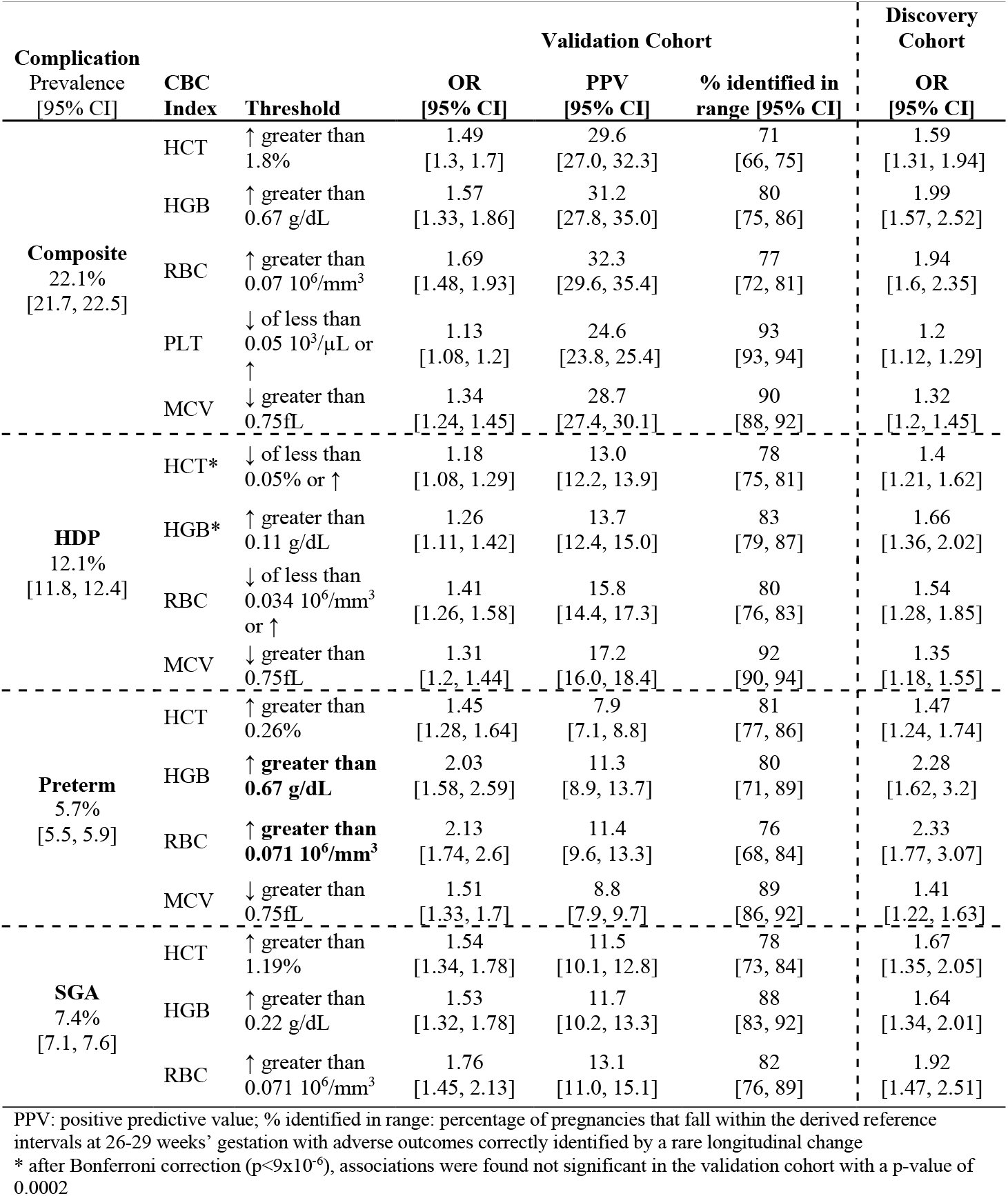
Pregnancies with rare longitudinal changes in some CBC indices between 7-14 and 26-29 weeks’ gestation have higher rates of complications. Listed are the OR and PPV in our validation cohort of the rare changes significantly associated with adverse outcomes (p<9×10^−6^, reflecting Bonferroni correction) and for which the positive predictive value (PPV) confidence interval did not overlap prevalence confidence intervals in our discovery cohort. Confidence intervals were obtained with bootstrapping. “% identified in range” denotes the percentage of pregnancies with adverse outcomes correctly identified with a rare behavior that fall within the 26-29 weeks’gestation reference interval.

Given these associations with the composite complication, we analyzed each complication separately and found significant associations between preterm birth and rare changes in HGB or RBC in the discovery; both associations were subsequently confirmed in the validation cohort. The preterm birth OR in the validation cohort was 2.0 for HGB [1.6, 2.6] p=2 x 10^−8^ and 2.1 for RBC [1.8, 2.6] p=9 x 10^−14^ for threshold increases of ≥ 0.67 (g/dL) for HGB or ≥ 0.07 (10^6^/mm^3^) in RBC (**Table 2**). PPVs for these rare changes were about twice the prevalence of preterm birth in the population. The subsequent preterm deliveries associated with these rare CBC changes included both spontaneous and medically indicated deliveries, and the proportion of each type of preterm delivery predicted was not distinguishable from the proportion in the full cohort. Most preterm deliveries associated with these rare changes had HGB and RBC within the 26-29 weeks’ gestation reference intervals and would not typically be identified based on that reference interval alone (**Table 2**). To explore the potential for early risk stratification, we analyzed the time intervals between detection of the rare behavior and preterm birth and found that it was > 7 weeks in the discovery cohort as well as in the validation cohort (7.9 weeks for HGB and 7.5 for RBC).

## Discussion

We derived gestational age-specific reference intervals for CBC indices and their changes during pregnancy in a large retrospective study of more than 95,000 pregnancies. These reference intervals help identify pregnancies in the study cohort with elevations (either absolute or relative to baseline) in some CBC indices that are at significantly greater risk of obstetric complications. In particular, elevations in red blood cell-related indices were associated with increased risk of subsequent HDP and preterm birth. We also found a doubling in risk of preterm birth for pregnancies with large increases in HGB or RBC between 7-14 and 26-29 weeks’ gestation. These findings highlight the opportunity to improve risk stratification for pregnant patients by systematically assessing longitudinal changes in routinely-available CBC indices.

The reference intervals identified in our cohort were noticeably wider than previously published trimester-specific intervals for RBC, MCV, MCH, and MCHC ^3,4,12,18,19,39^. These differences may in part reflect the smaller size of previous study cohorts^3,4,30^. Recent larger studies outside of the US found wider intervals for MCV and MCH, consistent with our results^5,40,41^. It is noteworthy that this study cohort’s HGB reference intervals includes values low enough to be considered anemia according to some current guidelines^32^, meaning that significant numbers of pregnancies have HGB levels that are normal according to a large cohort of uncomplicated pregnancies but will be labeled as anemic nevertheless. This difference may arise in part because current HGB thresholds for clinical management of anemia were in some cases derived from studies in which participants were iron-supplemented, potentially affecting the applicability to current pregnant individuals.^42^ The absence of association in our study between decreased red cell indices and adverse outcomes may be surprising and may been affected by the fact that individuals with low HGB or HCT values in our study were likely supplemented with iron based on current recommendation^43–45^.

The findings that elevations in HCT, HGB, and RBC were associated with higher risk of preeclampsia, HDP, SGA, and preterm birth are consistent with a smaller study which found that higher *first trimester* HGB concentrations were associated with an increased risk of gestational hypertension, preeclampsia, preterm birth, and SGA^46^. Three other smaller studies have linked higher HGB concentrations to stillbirths, HDP, and lower birth weight^8,47,48^. We speculate that high HGB values might be a sign of hemoconcentration or might be associated with abnormal placentation, both of which have been hypothesized to be involved in the pathophysiology of HDP, SGA, and preterm birth.^46,49^ Current guidelines on CBC interpretation during pregnancy focus primarily on low values, but our results suggest benefit in considering high values as indications for further evaluation. Although the frequency of elevated CBC indices is rare (<10%), this evaluation may nevertheless be cost-effective as CBCs are already routinely collected.

CBC changes during pregnancy were often dramatic, for instance with HGB and RBC for most pregnancies decreasing from baseline by more than twice what is expected for normal biological variation outside pregnancy.^33^ These changes are consistent with those reported in previous studies, which were smaller or cross-sectional^3,15,17–19,21–23^. By assessing the magnitude and direction of a change in a CBC index, each patient is used as their own baseline, an approach which has been shown to be informative outside pregnancy^7^. The reference intervals for CBC changes reported in this study can help identify deviations from health that are not likely to be detected if the absolute CBC results remain within reference. Indeed, most pregnancies with rare changes and subsequent complications had CBC indices within the 26-29 weeks’ reference interval.

We found that the rare increases in RBC or HGB between 7-14 and 26-29 weeks’ gestation were associated with increased risk of pregnancy complications, including preterm birth. The majority of pregnancies saw a decrease in RBC and HGB during this gestational period. These changes in RBC and HGB which presumably reflect healthy pregnancy physiology may be achieved by modulating red cell mass, plasma volume, or both. Plasma volume changes may be more important given current understanding of the importance of hemodilution^20,50,51^ and existing hypotheses related to hemoconcentration-related risk^46,49,52^. We also found an association between an extreme decrease in MCV and the composite outcome, and this decreased MCV may in some cases reflect iron deficiency that may have been undetected and untreated due to application of inappropriate anemia thresholds, but further study assessing iron status is necessary.

Our study has several strengths. First, we defined gestational-age-specific intervals in a cohort of patients in the US that is significantly larger than those typically used in prior studies that did not involve meta-analysis^3,4,30^. Second, we systematically analyzed longitudinal intra-pregnancy CBC dynamics and explored their use as prognostic factors for pregnancy complications. Third, our validated definitions of pregnancy complications were derived using rigorous methods and are likely more reliable than those based solely on administrative data^24^. Fourth, our key findings were validated in an out-of-sample cohort.

Our study also has limitations. First, it was retrospective and limited to pregnancies with prenatal care starting no later than 20 weeks’ of gestation, and while inclusion criteria were defined, incomplete data can lead to inappropriate inclusion of some patients with anemia or other pathologies. While our sensitivity analyses did not show major changes when anemic individuals were excluded, further investigations are warranted. Second, we evaluated longitudinal changes in pregnancy at three time points, but more frequent time sampling under controlled conditions could be more accurate. In particular, because the cohort was comprised of individuals who received obstetric care in our health system, not all participants had pre-pregnancy CBC data, which limited our sample size for examining longitudinal changes in CBC indices that occur between pre-pregnancy and the first trimester. Third, the method of ascertainment of HDP differed in the discovery cohort (blood pressure, laboratory, and labor and delivery record) from the validation cohort (ICD codes) which led to different prevalences of this outcome in the two cohorts and may have affected the validation of our results. Finally, the study population and chosen time windows might not accurately reflect obstetric practices outside of our health system.

In this study, we derive gestational-age-specific reference intervals for CBC indices and their longitudinal changes in a large US-based cohort. We use these references to find that elevations and rare longitudinal increases in red cell indices are associated with complications of pregnancy. Future work is needed to determine whether these findings can help understand the processes governing risk of obstetric complications and be prospectively integrated into clinical practice to improve outcomes.

## Data Availability

Data is available from the authors upon reasonable request and subject to approval from the Massachusetts General Brigham Institutional Review Board.

## Funding and Assistance

V.T, R.P. and J.M.H were supported by NIH DK123330. V.T, R.P, J.M.H and C.E.P were supported by NIH R01HD104756. K.J.G reports funding from NIH/NHLBI K08 HL146963. L.S. reports funding from NIH/NICHD 5K12HD103096-04. C.E.P. was supported by NIH/NIDDK K26DK138346. J.M. was supported by a T32 training grant for Endocrinology (T32DK007028-47S1). The MGH Maternal Health Cohort was supported by the MGH Claflin Distinguished Scholar Award and the MGH Physician-Scientist Development Award (to C.E.P.).

## Author contributions

V.T, R.P, J.M.H and C.E.P conceptualized, designed, and conducted the study as well as interpreted the results. V.T, R.P, K.J., S.H., B.F., C.M., J.M., C.B.C. performed data collection. V.T, R.P, J.M.H and C.E.P wrote the first draft of the manuscript, and all authors edited, reviewed, and approved the final version of the manuscript.

