## Supplementary for "Hematologic dynamics during pregnancy and their association with obstetric complications: a retrospective cohort study"

SUPPLEMENTARY TABLES .......... 1

**Supplementary methods**

Exclusion criteria for the discovery and validation cohorts

In the discovery cohort (**Figure 1**), pregnancies meeting the following criteria were excluded: late presentation to prenatal care (at >20 weeks' gestation), evidence of elevated blood pressure prior to 20 weeks' gestation, known chronic hypertension, stillbirth, missing documentation of birthweight or neonatal sex, no CBC data available at any point in pregnancy, delivery before 30 weeks' gestation (to prevent overlap between our exposure of a 26-29 weeks' gestation CBC and adverse outcomes of interest), non-singleton gestation, transfusion requirement during or within 8 weeks prior to pregnancy, or pre-existing diagnosis of immune thrombocytopenia purpura, hemolytic anemia, or a genetic blood disorder. More details on the hematological conditions are given below in Hematology related problems.

In the validation cohort, a total of 104,843 deliveries between 2016 and 2022 at any medical center within the Mass General Brigham healthcare system were considered. Pregnancies were excluded if they were known to be associated with a diagnosis of chronic hypertension (via ICD code), stillbirth or miscarriage, missing documentation of birthweight or neonatal sex, delivery before 30 weeks' gestation, non-singleton gestation, or pre-existing diagnosis of a blood disorder (ICD 10 codes D66 D67 D68 D69).

Hematology related problems

Patients diagnosed with any of the following blood disorders prior to pregnancy were excluded from the analysis in the discovery cohort. Diagnoses were ascertained from a clinical problem list in the obstetric record. Diagnostic exclusion criteria included:

- Immune/idopathic thrombocytopenia purpura (ITP)
- Alpha thalassemia trait and Hemoglobin H disease
- Beta thalassemia trait, beta thalassemia intermedia, and beta thalassemia major
- Sickle cell disease, including sickle-beta-zero thalassemia, sickle-beta-plus thalassemia, and hemoglobin SC disease. (Subjects with sickle trait were included in the analysis.)
- Hemoglobin E heterozygosity or homozygosity
- G6PD deficiency homozygosity (but not G6PD carriers)
- Hereditary spherocytosis
- Hereditary elliptocytosis
- Hereditary xerocytosis
- Pyruvate kinase deficiency
- Hemolytic anemia

**Complications of pregnancy in discovery cohort**

Hypertensive disorders of pregnancy

Pregnancies were considered affected by a hypertensive disorder of pregnancy (HDP), including gestational hypertension or preeclampsia, if there was no evidence of pre-existing chronic hypertension and at least one of the following was found in the medical record:

- Two or more systolic blood pressure (BP) readings >= 140 mmHG and/or diastolic BP readings >=90 mmHG greater than 4 hours apart after 20 weeks of gestation and up to one week postpartum;
  OR
- Recording of HDP as an indication for induction, indication for cesarean or complication of labor in the labor and delivery record;
  OR

- One elevated systolic BP >=140 mmHG and/or diastolic >=90 mmHG after 20 weeks of gestation with evidence of preeclampsia (see below).

Pregnancies were considered to be affected by preeclampsia if there was:
- Hypertensive disorder of pregnancy (as above)
  AND
- Evidence of preeclampsia as defined by one or more of the following:
  - 24 hour timed urine protein >=300 mg/day
    OR
  - Urine protein to creatinine ratio>=0.3 mg/mg Cr
    OR
  - Serum creatinine >=1.1 mg/dL
    OR
  - Platelets < 100 $10^9$/L
    OR
  - AST>40 U/L
    OR
  - ALT>40 U/L
    OR
  - Spot/dipstick urine protein reading >=2+ in the absence of timed urine or urine protein to creatinine ratio
    OR
  - ICD code for eclampsia

These HDP definitions were validated via blinded chart review by maternal fetal medicine specialists with an accuracy of 90%.

Small for gestational age

Newborns were considered small for gestational age (SGA) if their birthweight was less than the 10th percentile of babies of the same sex delivered at the same gestational week. See table below for the 10th percentile weight cutoffs used for the analyzed cohort defined in (Oken et al., 2003).

| | | Gestational age at delivery (weeks) | | | | | | | | | | | |
|---|---|---|---|---|---|---|---|---|---|---|---|---|---|
| | | **30** | **31** | **32** | **33** | **34** | **35** | **36** | **37** | **38** | **39** | **40** | **41** |
| **Weight (grams)** | **Female** | 965 | 1180 | 1390 | 1638 | 1872 | 2099 | 2299 | 2495 | 2694 | 2834 | 2919 | 2949 |
| | **Male** | 1044 | 1241 | 1475 | 1712 | 1957 | 2192 | 2410 | 2609 | 2807 | 2947 | 3029 | 3063 |

Transfusions

We considered transfusions of plasma, platelets, and red blood cells. Pregnancies for which individuals received a transfusion 8 weeks before or during pregnancy were excluded. Pregnancies with transfusions at delivery or within 8 weeks post-partum were included and classified as having had complications.

Complications of pregnancy in the validation cohort

For this cohort, small for gestational age and preterm adverse outcomes were defined as in the discovery cohort. Pregnancies with hypertensive disorders of pregnancy were identified using ICD codes (O13 O14.0 O14.9 O14.1 O14.2 O11) (Leonard et al., 2020). Preeclampsia as a subset of hypertensive disorders and transfusion were not defined in this dataset.

Routine pre-pregnancy CBCs

Routine pre-pregnancy CBCs were considered if the individual had no more than two complete blood count (CBC) tests in their medical record within the five years before their pregnancy separated by at least 6 months, and had no encounter marked with an ICD9 code related to pregnancy (V20-V29, 630-679), cancer (140-239), or infection (1-139) between 180 days prior the CBC and until the day of the pregnancy in consideration. A histogram of the PheCodes (Wu et al., 2019) for the included patients is available in **Figure S4**.

Estimation of body mass index (BMI) in discovery and validation cohorts

For each pregnancy included in the discovery cohort, we standardized prenatal weight/BMI to values at 12 weeks' gestation, interpolating or extrapolating if necessary, as described previously in (Selen et al., 2023).
For each pregnancy in the validation cohort, BMI was the pre-pregnancy BMI self-reported at the first trimester visit. Missing BMI (4% of considered pregnancies) were removed.

CBC measurements

CBCs were measured on Siemens Advia 2120 instruments until 2012 and on Sysmex XE-5000 instruments until 2016. Most CBC measurements performed between 2016 and 2022 were made on Sysmex XN-9000 instruments. All CBCs were measured in clinical laboratories that participated in proficiency testing programs.

CBC projection within gestational windows of consideration

In order to account for possible physiological variation within 7-14, 26-29, and 37-41 (pre-delivery) weeks' gestation, we estimated the CBC indices at the same fixed time points for each pregnancy: 9.5, 27.5, and 39.5 week. These values are the modes of distribution of visit times and reflect when pregnant individuals typically receive care at MGH (7-12 weeks', 26-29 weeks', 37-42 weeks'). We additionally included CBC indices between 12-14 weeks' gestation for analysis to include individuals who discovered their pregnancies at a slightly later gestational age. These midpoints were used in the linear mixed-effects model described below. We estimated the change within those weeks across patients as illustrated below for RBC and RDW.

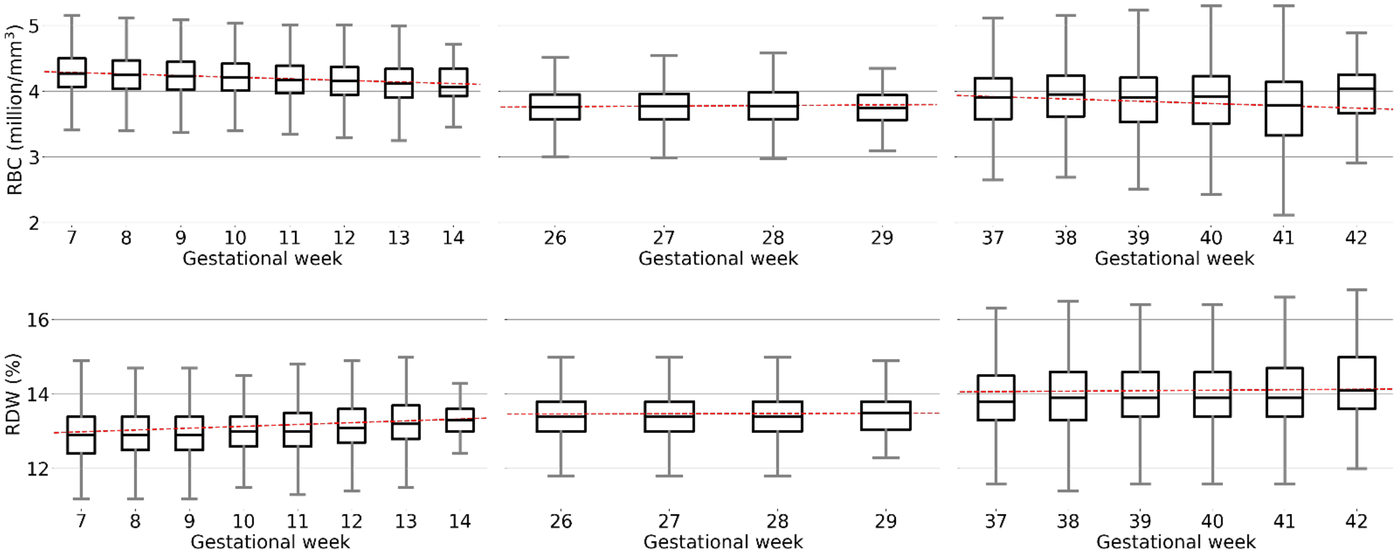

We inferred a projection coefficient using a linear mixed-effects model adjusting for BMI at 12 weeks' gestation and maternal age. We also add random intercepts for individual pregnancies to account for physiological variation between pregnancies in a single patient. If we consider the example of RBC changes in the following time interval $t = [7, 14]$, then the model is formally written as $RBC_{it} = \beta_0 + \beta_1^{RBC,t} GA_{it} + \beta_2 BMI_i + \beta_3 Age_i + b_{0i}$ where $i$ denotes the pregnancy and $GA_{it}$ denotes the gestational age that falls within the interval [7, 14]. $b_{0i}$ is the random intercept for each pregnancy. The coefficient used for the final projection is $\beta_1$, and the list of coefficients for the different CBC indices as well as time intervals is reported below. To obtain the averaged and projected $RBC_{i,7-14}$ in the interval 7-14 weeks to 9.5 weeks for a pregnancy $i$, we compute

$$RBC_{i,7-14} = \frac{1}{n_{i,7-14}} \sum_{j=1}^{n_{i,7-14}} \left[RBC_{i,j} + \beta_1^{RBC,7-14} \cdot (9.5 - GA_{i,j})\right]$$

where $n_{i,7-14}$ is the number of CBCs measured in the considered time period for pregnancy $i$, $\beta_1^{RBC,7-14}$ is the projection coefficient estimated above, and $GA_{i,j}$ is the specific gestational age for the $j$-th measured $RBC_{i,j}$.

| | Projection coefficients $\beta$ (Std.Err) P=p-value | | |
|---|---|---|---|
| CBC index | 7-14 weeks | 26-29 weeks | 37-42 weeks |
| Hematocrit | -0.249 (0.008) p<0.01 | -0.106 (0.017) p<0.01 | -0.311 (0.017) p<0.01 |
| Hemoglobin | -0.058 (0.003) p<0.01 | -0.025 (0.006) p<0.01 | -0.109 (0.006) p<0.01 |
| White blood cell count | -0.02 (0.007) p=0.01 | -0.011 (0.016) p=0.48 | 0.207 (0.014) p<0.01 |
| Red blood cell count | -0.023 (0.001) p<0.01 | 0.004 (0.002) p=0.04 | -0.042 (0.002) p<0.01 |
| Platelet count | -2.519 (0.174) p<0.01 | -0.503 (0.393) p=0.2 | 1.797 (0.255) p<0.01 |
| Mean Corpuscular Volume | -0.049 (0.01) p<0.01 | -0.305 (0.029) p<0.01 | 0.158 (0.012) p<0.01 |
| Mean Corpuscular Hemoglobin | 0.033 (0.004) p<0.01 | -0.082 (0.011) p<0.01 | -0.066 (0.005) p<0.01 |
| Red blood cell Distribution Width | 0.058 (0.003) p<0.01 | 0.006 (0.005) p=0.25 | 0.021 (0.003) p<0.01 |
| Mean Corpuscular Hemoglobin Concentration | 0.073 (0.004) p<0.01 | 0.034 (0.008) p<0.01 | -0.047 (0.005) p<0.01 |

Beta coefficient derivation

Intra-patient trends in CBC indices over the course of prenatal visits were analyzed using a linear mixed-effects model adjusted for age and BMI at 12 weeks' gestation. Random effects were added for the individual pregnancy to account for physiological variation at pre-delivery, for individual patients, for year of delivery, for parity, and for different prenatal sites of care to account for machine calibration biases. If we consider the example of RBC dynamics between a [7,14] week visit and a [26,29] week visit, then the model is formally written as:

$$RBC_i = \beta_0 + \beta_1 Visit_i + \beta_2 BMI_i + \beta_3 Age_i + b_{0i} + b_{0j} + b_{0k} + b_{0y} + b_{0z}$$

where $i$ denotes the pregnancy, $j$ denotes the prenatal care location, $k$ denotes the year, $y$ denotes parity, and $z$ denotes the individual. $\beta_1 Visit_i$ is the contribution that the distance between two successive prenatal visits, or pre-pregnancy to the first prenatal visit, contributes to the change in that marker. To more uniformly evaluate the contribution of gestational age, all CBC indices from a given pregnancy were projected to the mode week of the time frame, as described above in CBC projection within gestational windows of consideration.

Odds ratio derivation

Odds ratios were obtained using generalized estimating equations (GEE) for logistic regression adjusted for BMI at 12 weeks' gestation, age of the individual, insurance, parity, and race/ethnicity. Clustering was at the level of the individual and an exchangeable correlation structure was used. The model for the mean is as follows:

$$logit(E[Y_i]) = \beta_0 + \beta_1 Indicator_i + \beta_2 BMI_i + \beta_3 Age_i + \beta_4 Insurance_i + \beta_5 Parity_i + \beta_6 Race/ethnicity_i$$

where $E[Y_i]$ denotes the expected value, or probability, of an adverse outcome of interest (small for gestational age, preeclampsia, hypertensive disorder of pregnancy, preterm delivery, transfusion at or after delivery) or a composite of HDP, preterm delivery, and SGA, and $i$ denotes the pregnancy. $Insurance$ and $Race/ethnicity$ categories are presented in **Table 1**. Race/ethnicity is included in this analysis to capture disparities in adverse pregnancy outcomes among self-reported racial groups and not to imply any biological mechanism underlying these disparities. $Indicator$ is a binary indicator variable marking whether the individual fell within or outside of this study's reference intervals or those in the literature, or met definition of a rare dynamic (see Rare dynamic definition) depending on the test at hand. For reference interval tests, we considered whether CBC values in a pregnancy fell outside of the reference interval for that index at 26-29 weeks, in a one-sided (above or below) and two-sided (above and below) fashion. Rare behavior tests are described in Rare dynamic definition. The package geepack in R was used to estimate regression coefficients (Højsgaard et al., 2006; Yan, 2002; Yan & Fine, 2004). P-values were corrected with a Bonferroni correction, the level of which was test specific and is reported in appropriate figure and table captions (**Figure 3**, **Table 2** and **Tables S6-8**). For this analysis we excluded pregnancies with a HDP diagnosis before 29 weeks in the discovery cohort as determined by an elevated blood pressure.

### Calculation of biological variation

Biological variation is used as a benchmark for changes in CBC results. Intra-person biological variation is estimated for each index by the European Federation of Clinical Chemistry and Laboratory Medicine (EFLM) Working Group on Biological Variation using meta-analyses of high quality studies, ascertained through the Biological Variation Data Critical Appraisal Checklist, to describe the variability of laboratory indices within individuals. The biological variation was provided as a coefficient of variation which was used to calculate values representing 2 standard deviations from the mean of each CBC index in our discovery cohort. The column Biological variation is the intra-patient coefficient of variation for that marker as available from (Aarsand et al., n.d.) at the link https://biologicalvariation.eu. The column Mean is the value used to estimate the standard deviation and is calculated as the mean at the pre-pregnancy point across patients with no adverse events. The column 2STD cutoff is two times the biological variation multiplied by the mean of the pre-pregnancy timepoint, and serves as the cutoff to which we compare subsequent dynamics of the CBC indices (see **Figure 2 and Table 2**).

| **CBC index** | **Biological variation (%)** | **Mean** | **2STD cutoff** |
|---|---|---|---|
| Hematocrit (%) | 2.8 | 39 | 2.2 |
| Hemoglobin | 1.6 | 13.2 | 0.4 |
| White blood cell count | 10.8 | 7.3 | 1.6 |
| Red blood cell count | 2.6 | 4.4 | 0.2 |
| Platelet count | 7.5 | 271 | 40.7 |
| Mean Corpuscular Volume | 0.8 | 88 | 1.4 |
| Mean Corpuscular Hemoglobin | 0.9 | 30 | 0.5 |
| Red blood cell Distribution Width (%) | 1.6 | 12.8 | 0.4 |
| Mean Corpuscular Hemoglobin Concentration | 1 | 34.2 | 0.7 |

### Rare dynamic definition

We analyzed data from three time intervals (pre-pregnancy to 7-14 weeks' gestation, 7-14 weeks' to 26-29 weeks' gestation, and 26-29 weeks' gestation to pre-delivery). In the main analysis we focused on the time interval between 7-14 weeks and 26-29 as the most unbiased and practical time point that for detection of pregnancies with adverse outcomes before delivery. As CBCs among young people capable of pregnancy are not routinely indicated as screening tests outside of pregnancy, individuals with pre-pregnancy CBCs may have selection bias and may not reflect indices in the general population. Additionally, 64% of analyzed pregnancies have a CBC at both 7-14 weeks' and 26-29 weeks' gestation, compared to just 9% and 10% respectively for a CBC at both pre-pregnancy and 7-14 weeks'. Analyzing changes between 26-29 weeks' gestation and pre-delivery would not allow for timely prediction of complications. Thus, we focused on the interval between 7-14 weeks' and 26-29 weeks' gestation for prediction of adverse outcomes.

For this interval between 7-14 weeks' and 26-29 weeks' gestation we compute the change in the CBC indices (delta) for each pregnancy. We considered a delta to be stable if absolute value of the change is less than $2x$ the biological variation for that index, increasing if it is greater than $2x$ the biological variation, and decreasing if it is less $-2x$ of the biological variation. Biological variation for each marker was derived from the EFLM Biological Variation Database (Aarsand et al., n.d.). We reported the estimated standard deviations above in Calculation of biological variation. The prevalence of observed CBC index dynamics was calculated from the percentage of patients with positive, non-changing, or negative deltas. The directionality of the rare behavior was decided based on the least likely of the three types of change between 7-14 and 26-29 weeks' gestation (see **Table S5**). After directionality was chosen, we tested 100 thresholds linearly spaced between minimum and maximum delta values for each index, and selected the thresholds yielding the highest positive predictive value (PPV) and a significant odds ratio (OR). We focused on PPV because it helps assess a marker's potential clinical significance, which typically requires that the PPV significantly exceed the prevalence of the outcome being predicted. Significance of the OR was decided by comparing p-values with a Bonferroni corrected threshold, $\alpha$ =0.05/(100 thresholds*9 markers*6 adverse outcomes). See **Table 2** in main text for rare dynamics significantly associated with adverse outcomes, and **Table S8** for a summary of other adverse outcomes and indices.

### Supplementary figures

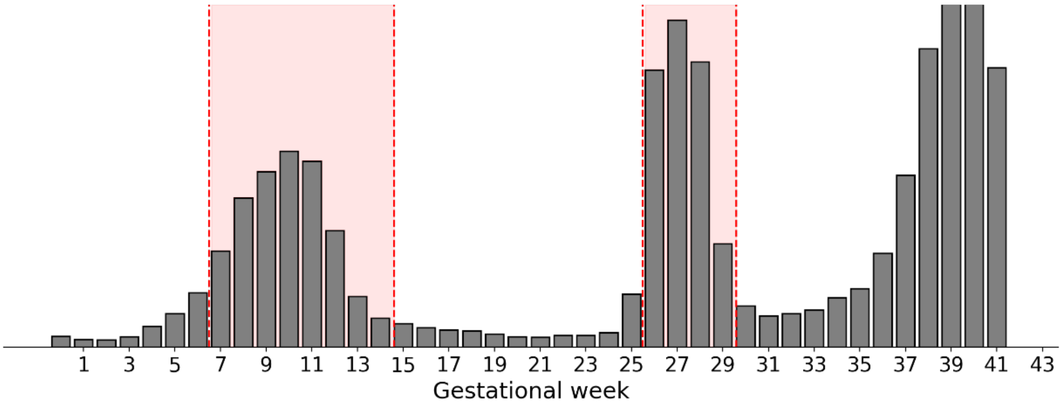

**Figure S1**. Frequency of available CBCs by gestational week in discovery cohort pregnancies. Shaded red areas indicate chosen windows for which we considered CBCs in gestation (7-14 weeks and 26-29 weeks). The pre-delivery timepoint is not shaded as it is individualized and fell between 30 to 41 weeks gestation.

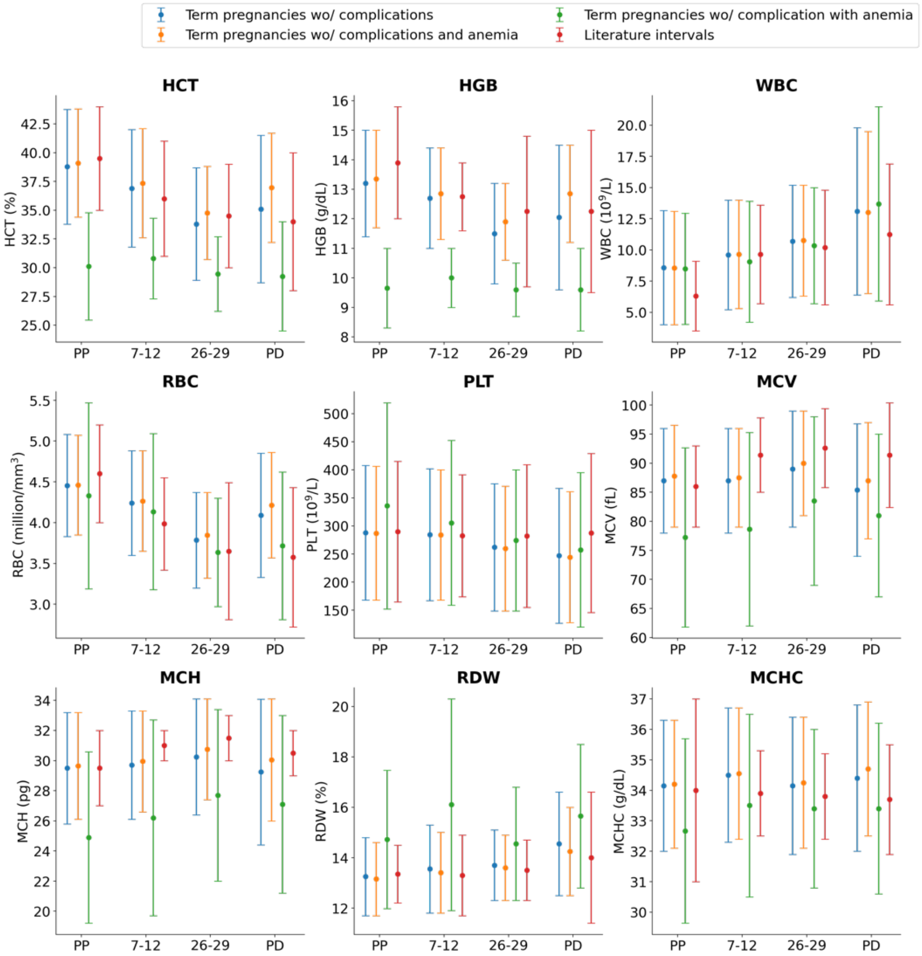

**Figure S2**. **Sensitivity analysis of the effects of including patients with a diagnosis of anemia on reference interval determination**. The figure compares gestational-age-specific intervals for all term pregnancies without complications (blue error bars), pregnancies with no diagnosis of anemia (<11 g/dL HGB at PP, 7-14 and PD, and <10.5 g/dL at 26-29 weeks) (orange error bars), pregnancies considered anemic (green error bars), and literature trimester-specific intervals (red error bars), all in the discovery cohort. Up to 15% of pregnancies met criteria for anemia at each timepoint.

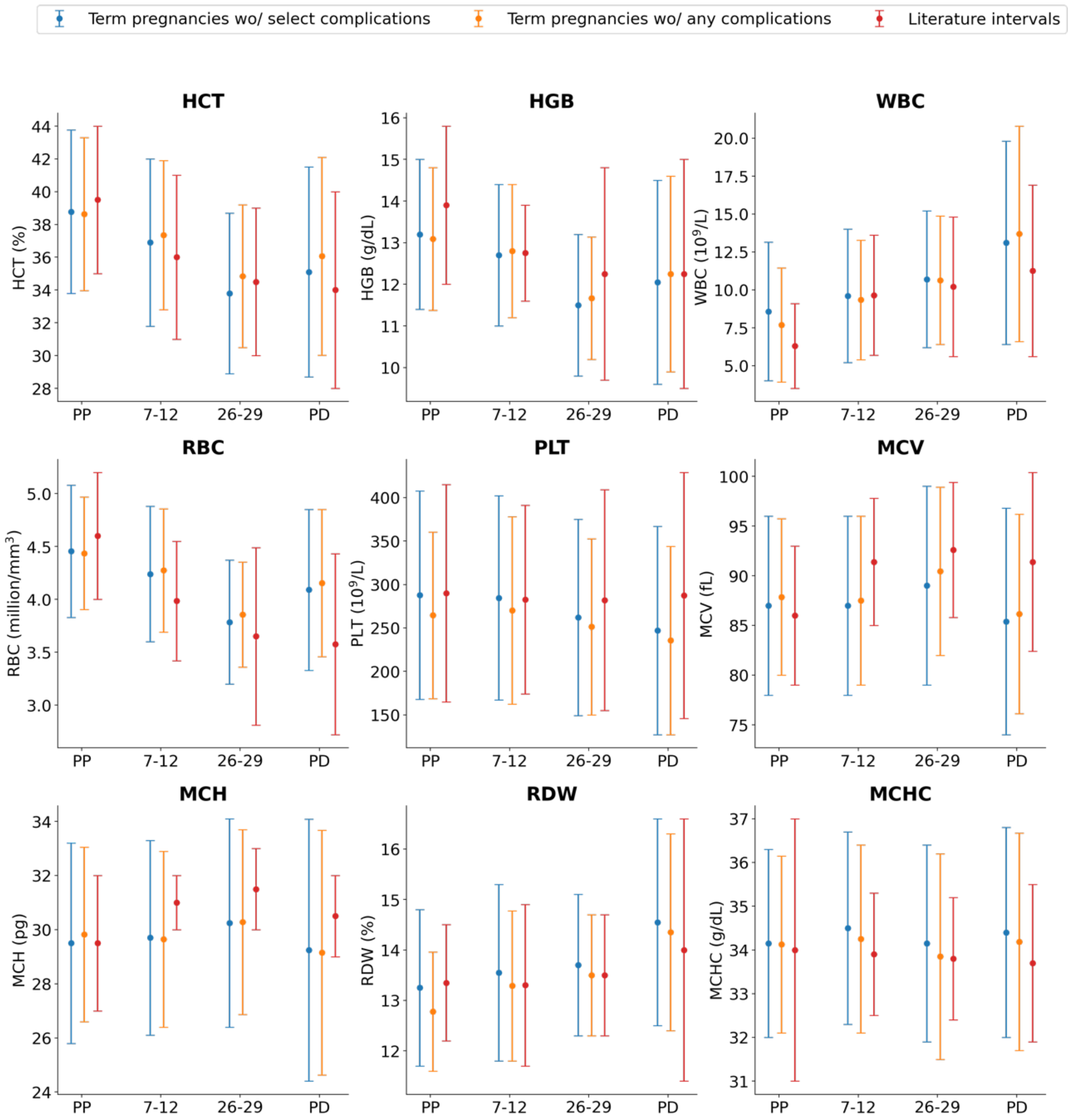

**Figure S3**. **Sensitivity analysis of the effects of including patients with chronic or pregnancy-related conditions on reference intervals**. Comparison of gestational-age-specific intervals for term pregnancies without complications in discovery, pregnancies in individuals without chronic or pregnancy-related conditions (N=1460) in discovery and literature trimester-specific intervals.

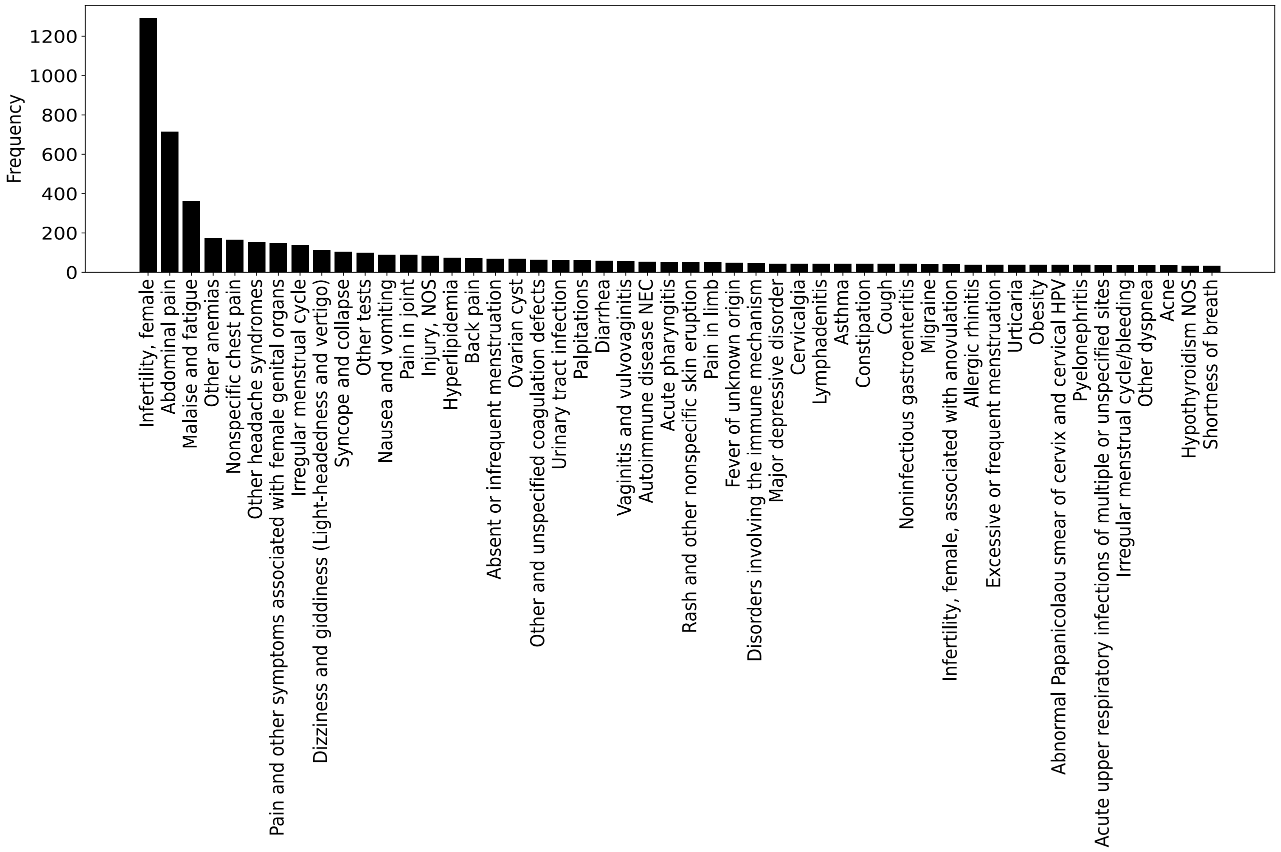

**Figure S4.** Histogram of 50 most frequent PheCodes for visits corresponding to the pre-pregnancy CBCs considered. Considered pre-pregnancy CBCs are described in Routine pre-pregnancy CBCs.

**Supplementary Tables**

**Table S1**. Tabular version of reference intervals derived from our cohort and literature intervals.

| | Pregnancy without complications | | | | Literature reference ranges | | | | |
|---|---|---|---|---|---|---|---|---|---|
| | Pre-pregnancy | 7-14 weeks | 26-29 weeks | Pre-Delivery | Non-pregnant | First Trimester | Second trimester | Third trimester | Ref. |
| Hematocrit (%) | [33.8, 43.8] Mean: 38.8 | [31.8, 42.0] Mean: 36.9 | [28.9, 38.7] Mean: 33.8 | [28.7, 41.5] Mean: 35.6 | [35, 44] | [31, 41] | [30, 39] | [28, 40] | [1] |
| Hemoglobin (g/dL) | [4.0, 13.2] Mean: 7.3 | [5.2, 14.0] Mean: 9.1 | [6.2, 15.2] Mean: 10.1 | [6.4, 19.8] Mean: 11.7 | [12, 15.8] | [11.6, 13.9] | [9.7, 14.8] | [9.5, 15] | [1] |
| White blood cell count ($10^9$/L) | [168.0, 407.5] Mean: 271.3 | [167.0, 402.0] Mean: 269.3 | [149.0, 375.0] Mean: 245.7 | [127.0, 367.0] Mean: 227.8 | [3.5, 9.1] | [5.7, 13.6] | [5.6, 14.8] | [5.6, 16.9] | [1] |
| Red blood cell count ($10^6$/mm$^3$) | [25.8, 33.2] Mean: 30.1 | [26.1, 33.3] Mean: 30.3 | [26.4, 34.1] Mean: 30.8 | [24.4, 34.1] Mean: 30.1 | [4, 5.2] | [3.4, 4.6] | [2.8, 4.5] | [2.7, 4.4] | [1] |
| Platelet count ($10^9$/L) | [32.0, 36.3] Mean: 34.2 | [32.3, 36.7] Mean: 34.5 | [31.9, 36.4] Mean: 34.2 | [32.0, 36.8] Mean: 34.6 | [165, 415] | [174, 391] | [155, 409] | [146, 429] | [1] |
| Mean Corpuscular Volume (fL) | [11.4, 15.0] Mean: 13.3 | [11.0, 14.4] Mean: 12.7 | [9.8, 13.2] Mean: 11.6 | [9.6, 14.5] Mean: 12.3 | [79, 93] | [85, 98] | [86, 99] | [82, 100] | [1] |
| Mean Corpuscular Hemoglobin (pg) | [3.8, 5.1] Mean: 4.4 | [3.6, 4.9] Mean: 4.2 | [3.2, 4.4] Mean: 3.8 | [3.3, 4.8] Mean: 4.1 | [27, 32] | [30, 32] | [30, 33] | [29, 32] | [1] |
| Red blood cell Distribution Width (%) | [78.0, 96.0] Mean: 88.1 | [78.0, 96.0] Mean: 87.8 | [79.0, 99.0] Mean: 90.2 | [74.0, 96.8] Mean: 87.2 | <14.5 | [11.7, 14.9] | [12.3, 14.7] | [11.4, 16.6] | [1][2] |
| Mean Corpuscular Hemoglobin Concentration (g/dL) | [11.7, 14.8] Mean: 12.8 | [11.8, 15.3] Mean: 13.1 | [12.3, 15.1] Mean: 13.5 | [12.5, 16.6] Mean: 14.0 | [31, 37] | [32.5, 35.3] | [32.4, 35.2] | [31.9, 35.5] | [2][3] |

**Table S2.** The number of total pregnancies and the number of pregnancies with various complications in both the discovery and validation cohorts. Pregnancies with a first elevated blood pressure before 29 weeks' gestation are excluded from the group with a CBC at 26-29 weeks' gestation and the group with CBCs at both 7-14 and 26-29 weeks' gestation.

| Complication | Discovery | | | Validation |
|---|---|---|---|---|
| | Pregnancies with CBC at any time point | Pregnancies with CBC at 26-29 weeks | Pregnancies with CBCs at both 7-14 and 26-29 weeks | Pregnancies with CBCs at both 7-14 and 26-29 weeks |
| All pregnancies | 45,992 (100%) | 33,862 (100%) | 29,162 (100%) | 50,603 (100%) |
| All complications | 8,283 (18%) | 5,921 (17%) | 5,037 (17%) | N/A |
| Composite | 8,061 (18%) | 5,763 (17%) | 4,907 (17%) | 11,175 (22%) |
| Pre-eclampsia | 1,573 (3%) | 1,052 (3%) | 898 (3%) | N/A |
| HDP (Gestational hypertension and pre-eclampsia) | 3,219 (7%) | 2,173 (6%) | 1,862 (6%) | 6,128 (12%) |
| Small for gestational age | 3,253 (7%) | 2,356 (7%) | 1,980 (7%) | 3,731 (7%) |
| Preterm birth between 30 and 37 weeks' gestation | 2,587 (6%) | 1,872 (6%) | 1,585 (5%) | 2901 (6) |
| Peripartum transfusion | 319 (0.7%) | 234 (0.7%) | 197 (0.7%) | N/A |

**Table S3.** Demographics of out-of-sample validation cohort of MGB deliveries between 2016 and 2022.

| Variable | | Value N(%) | Value N(%) |
|---|---|---|---|
| Pregnancies N (%) | | 39428 (100) | 11175 (100) |
| Age Mean (SD) | | 33.0 (4) | 33.0 (5) |
| Insurance Status N (%) | | | |
| | Public | 4847 (12) | 1584 (14) |
| | Private | 34219 (87) | 9496 (85) |
| | None | 362 (1) | 95 (1) |
| Self-reported race N (%) | | | |
| | Asian | 4456 (11) | 1248 (11) |
| | Black | 2304 (6) | 910 (8) |
| | Latina | 3265 (8) | 1006 (9) |
| | White | 28390 (72) | 7727 (69) |
| | None of the above | 1013 (2) | 284 (2) |
| Nulliparous N (%) | | 17354 (44) | 6589 (59) |
| Gestational age at delivery Mean (SD) | | 40.0 (1) | 38.0 (2) |
| Pregravida BMI Mean (SD) | | 25.2 (5.3) | 26.6 (6.3) |
| Time of CBC Measurement | | | |
| | 26-29 weeks' gestation | 39428 (100) | 11175 (100) |
| | 7/14 & 26/29 weeks' gestation | 39428 (100) | 11175 (100) |

**Table S4. Beta coefficients of CBC dynamics in pregnancy**. Beta coefficients measure the effect of gestational age on CBC values and were estimated on the subset of pregnancies with all four time points available (N=2,610). Random effects were added for the individual pregnancy, the individual, parity, OB-GYN site of care, and for year of delivery. The p-value threshold with Bonferroni correction for multiple testing was p=0.002.

| | PP - 7-14 (N=2791) | | 7-14 - 26-29 (N=2791) | | 26-29 - PD (N=2791) | |
|---|---|---|---|---|---|---|
| | β (Std. Err) | P | β (Std. Err) | P | β (Std. Err) | P |
| Hematocrit (%) | -1.84 (0.05) | 1.9e-258 | -3.19 (0.04) | 0 | 2.15 (0.06) | 2.6e-258 |
| Hemoglobin (g/dL) | -0.53 (0.02) | 5.6e-207 | -1.18 (0.01) | 0 | 0.91 (0.02) | 0 |
| White blood cell count ($10^9$/L) | 2.11 (0.05) | 0 | 0.97 (0.04) | 1.5e-131 | 1.94 (0.06) | 1.0e-182 |
| Red blood cell count ($10^6$/$mm^3$) | -0.2 (0.01) | 6.4e-280 | -0.45 (0) | 0 | 0.35 (0.01) | 0 |
| Platelet count ($10^9$/L) | 0.88 (0.8) | 0.3 | -25.17 (0.7) | 5.2e-234 | -22.98 (0.72) | 1.3e-190 |
| Mean Corpuscular Volume (fL) | -0.14 (0.05) | 3.4e-3 | 2.22 (0.05) | 0 | -2.49 (0.05) | 0 |
| Mean Corpuscular Hemoglobin (pg) | 0.2 (0.02) | 2.0e-23 | 0.54 (0.02) | 2.4e-159 | -0.38 (0.02) | 1.1e-65 |
| Red blood cell Distribution Width (%) | 0.17 (0.01) | 1.8e-30 | 0.41 (0.01) | 7.23e-163 | 0.42 (0.02) | 4.2e-135 |
| Mean Corpuscular Hemoglobin Concentration (g/dL) | 0.28 (0.02) | 5.1e-36 | -0.24 (0.02) | 1.0e-36 | 0.49 (0.02) | 2.3eE-133 |

**Table S5. Rare CBC changes (<5%) during pregnancy occur for red blood cell count, hematocrit, hemoglobin, platelet count, and mean corpuscular volume.** Percentages of pregnancies with decreasing, stable, or increasing CBC indices between subsequent timepoints for analysis of common and uncommon behavior in blood dynamics during pregnancy are shown. We considered a difference between two timepoints to be stable if the absolute value of the change was less than the biological variation for that index, increasing if it was positive and greater than biological variation, and decreasing if it was negative and greater in magnitude than biological variation (see **Methods – Statistical analysis** for further details). Additionally, percentage of pregnancies with stable (within biological variation) indices throughout gestation are reported in the last column. The cases changes exhibited by fewer than 5% of pregnancies are bolded.

| | Change 7-14 weeks' gestation - Pre-Pregnancy | | | Change 26-29 weeks' gestation - 7-14 weeks' gestation | | | Change Pre-delivery - 26-29 weeks' gestation | | | Change throughout pregnancy* |
|---|---|---|---|---|---|---|---|---|---|---|
| | Decrease | Stable | Increase | Decrease | Stable | Increase | Decrease | Stable | Increase | Stable |
| Hematocrit (%) | 47.8 | 46.8 | 5.4 | 65.5 | 33.3 | **1.3** | 7.6 | 46.6 | 45.8 | 6 |
| Hemoglobin (g/dL) | 56.4 | 33.4 | 10.2 | 82.3 | 15.7 | **2** | 10.2 | 24.1 | 65.7 | **1** |
| White blood cell count ($10^9$/L) | 5.4 | 33.8 | 60.8 | 8.4 | 54.4 | 37.3 | 11.1 | 45.5 | 43.4 | 7 |
| Red blood cell count ($10^6$/mm$^3$) | 48.5 | 46.6 | **4.9** | 81.2 | 18.4 | **0.4** | **4.4** | 31.6 | 64 | **3** |
| Platelet count ($10^9$/L) | 15 | 69.8 | 15.1 | 30.6 | 65.5 | **4** | 27.3 | 68.4 | **4.3** | 32 |
| Mean Corpuscular Volume (fL) | 28.4 | 49 | 22.5 | 5.7 | 31.4 | 63 | 63.1 | 32.7 | **4.2** | 7 |
| Mean Corpuscular Hemoglobin (pg) | 21.3 | 44 | 34.7 | 13.4 | 38.2 | 48.4 | 40.5 | 42.5 | 17 | 9 |
| Red blood cell Distribution Width (%) | 13.8 | 54.1 | 32.1 | 10.1 | 42.6 | 47.3 | 10.6 | 45.5 | 43.9 | 10 |
| Mean Corpuscular Hemoglobin Concentration (g/dL) | 19 | 41.6 | 39.3 | 36 | 46.3 | 17.7 | 15 | 43.9 | 41.1 | 9 |

* At each time point we considered the pregnancy stable if the change compared to previous time point was within biological variation. The percentage showed is the percentage of pregnancies that are stable at all time delta throughout pregnancy.

**Table S6. Beta coefficients are consistent when inferred on all available pregnancies with CBCs for at least two consecutive time points.** Beta coefficients were inferred on the subset of pregnancies for which at least one consecutive pair of time points was available (N=28,450) and compared to the beta coefficients in the main analysis. P-values were corrected for multiple testing with Bonferroni correction so that p<0.002 was considered significant. In bold are the p-values that denote a difference between the sensitivity analysis result and the main analysis, though all absolute differences in beta coefficients between sensitivity and main analysis fall within 2x biological variation.

| | PP -> 7-14 (N=3333) | | | 7-14 -> 26-29 (N=24125) | | | 26-29 -> PD (N=27795) | | |
|---|---|---|---|---|---|---|---|---|---|
| | $\beta$ (Std. Err) | P | P zeta test | $\beta$ (Std. Err) | P | P zeta test | $\beta$ (Std. Err) | P | P zeta test |
| Hematocrit (%) | -1.72 (0.04) | 1.6e-267 | 0.13 | -3.28 (0.01) | 0.0 | 0.30 | 1.92 (0.02) | 0 | 0.05 |
| Hemoglobin (g/dL) | -0.49 (0.01) | 6.6e-189 | **7.4e-06** | -1.21 (0.0) | 0.0 | 0.07 | 0.81 (0.01) | 0 | **6.4e-08** |
| White blood cell count ($10^9$/L) | 1.8 (0.04) | 0.0 | 0.0063 | 0.98 (0.01) | 0.0 | 0.47 | 1.57 (0.02) | 0 | **0.001980003** |
| Red blood cell count ($10^6$/mm$^3$) | -0.18 (0.01) | 3.6e-200 | **3.6e-54** | -0.47 (0.0) | 0.0 | **1.9e-09** | 0.34 (0.0) | 0 | **1.5e-10** |
| Platelet count ($10^9$/L) | 0.21 (0.67) | 0.89 | 0.14 | -26.16 (0.23) | 0.0 | 0.48 | -20.56 (0.22) | 0 | 0.44 |
| Mean Corpuscular Volume (fL) | -0.17 (0.04) | 0.0002 | 0.0291 | 2.31 (0.02) | 0.0 | 0.27 | -2.59 (0.02) | 0 | 0.25 |
| Mean Corpuscular Hemoglobin (pg) | 0.19 (0.02) | 1.8e-29 | 0.38 | 0.56 (0.01) | 0.0 | 0.11 | -0.49 (0.01) | 0 | **2.2e-29** |
| Red blood cell Distribution Width (%) | 0.22 (0.02) | 9.7e-42 | **5.9e-31** | 0.41 (0.0) | 0.0 | 0.31 | 0.48 (0.01) | 0 | **3.4e-16** |
| Mean Corpuscular Hemoglobin Concentration (g/dL) | 0.28 (0.02) | 9.7e-47 | 0.17 | -0.24 (0.01) | 6.2e-306 | 0.13 | 0.41 (0.01) | 0 | **5.3e-16** |

**Table S7. Elevated CBC index values are associated with composite outcome**. Pregnancies with a CBC at 26-29 weeks' gestation were considered (N=34,159), and those with abnormal tests for HDP or preeclampsia diagnosis before 29 weeks' gestation were excluded (N=254). The odds ratios of developing examined complications (composite of HDP, SGA, and preterm delivery; each of those adverse events individually; and need for transfusion at or after delivery) are shown in table for a CBC index value above, outside, or below the study intervals at 26-29 weeks' gestation. Significant odds ratios are marked with a 1 in the corresponding column. Prevalence, positive predictive value (PPV), and negative predictive value (NPV) are also reported. Significance was evaluated with a Bonferroni corrected p-value of 0.0002.

| Composite (All Complications Except Transfusion) | | | | |
|---|---|---|---|---|
| **Marker** | **Direction** | **Significant** | **O.R [95% CI], p** | **Prevalence/PPV/NPV** |
| HCT | Above range | 1 | 1.42 [1.21, 1.67] p=1.8e-05 | 0.17 / 0.23 / 0.83 |
| | Two-sided | 0 | 1.2 [1.06, 1.37] p=0.0047574 | 0.17 / 0.2 / 0.83 |
| | Below range | 0 | 0.94 [0.76, 1.16] p=0.5557298 | 0.17 / 0.16 / 0.83 |
| HGB | Above range | 1 | 1.65 [1.42, 1.92] p=1.4e-10 | 0.17 / 0.26 / 0.83 |
| | Two-sided | 1 | 1.34 [1.18, 1.52] p=4.4e-06 | 0.17 / 0.22 / 0.83 |
| | Below range | 0 | 0.94 [0.76, 1.16] p=0.5701551 | 0.17 / 0.16 / 0.83 |
| RBC | Above range | 1 | 1.61 [1.37, 1.88] p=3.9e-09 | 0.17 / 0.26 / 0.83 |
| | Two-sided | 0 | 1.25 [1.1, 1.42] p=0.0004937 | 0.17 / 0.21 / 0.83 |
| | Below range | 0 | 0.85 [0.69, 1.05] p=0.1377252 | 0.17 / 0.15 / 0.83 |
| WBC | Above range | 0 | 1.32 [1.11, 1.57] p=0.0015407 | 0.17 / 0.24 / 0.83 |
| | Two-sided | 0 | 1.15 [1.01, 1.31] p=0.0354735 | 0.17 / 0.19 / 0.83 |
| | Below range | 0 | 0.95 [0.78, 1.16] p=0.6230269 | 0.17 / 0.15 / 0.83 |
| PLT | Above range | 0 | 1.25 [1.06, 1.48] p=0.0096452 | 0.17 / 0.22 / 0.83 |
| | Two-sided | 0 | 1.18 [1.04, 1.34] p=0.0105382 | 0.17 / 0.19 / 0.83 |
| | Below range | 0 | 1.09 [0.9, 1.31] p=0.3828963 | 0.17 / 0.17 / 0.83 |
| MCV | Above range | 0 | 0.84 [0.67, 1.05] p=0.1299678 | 0.17 / 0.15 / 0.83 |
| | Two-sided | 0 | 1.05 [0.91, 1.2] p=0.5377823 | 0.17 / 0.18 / 0.83 |
| | Below range | 0 | 1.21 [1.01, 1.45] p=0.0342684 | 0.17 / 0.21 / 0.83 |
| MCH | Above range | 0 | 0.92 [0.75, 1.12] p=0.3964878 | 0.17 / 0.15 / 0.83 |
| | Two-sided | 0 | 1.01 [0.88, 1.16] p=0.8553496 | 0.17 / 0.17 / 0.83 |
| | Below range | 0 | 1.1 [0.91, 1.33] p=0.3054957 | 0.17 / 0.19 / 0.83 |

| | | | | |
|---|---|---|---|---|
| RDW | Above range | 0 | 0.95 [0.78, 1.15] p=0.5874473 | 0.17 / 0.18 / 0.83 |
| | Two-sided | 0 | 0.99 [0.86, 1.15] p=0.9209756 | 0.17 / 0.18 / 0.83 |
| | Below range | 0 | 1.05 [0.85, 1.31] p=0.6405973 | 0.17 / 0.18 / 0.83 |
| MCHC | Above range | 0 | 1.07 [0.89, 1.3] p=0.4624188 | 0.17 / 0.17 / 0.83 |
| | Two-sided | 0 | 1.09 [0.96, 1.25] p=0.1952262 | 0.17 / 0.18 / 0.83 |
| | Below range | 0 | 1.11 [0.92, 1.33] p=0.2856466 | 0.17 / 0.19 / 0.83 |

**Table S8. Elevated CBC index values are associated with preterm**. Pregnancies with a CBC at 26-29 weeks' gestation were considered (N=34,159). The odds ratios of developing examined complications are shown in table for a CBC index value above, outside, or below the study intervals at 26-29 weeks' gestation. Significant odds ratios are marked with a 1 in the corresponding column. Prevalence, positive predictive value (PPV), and negative predictive value (NPV) are also reported. Significance was evaluated with a Bonferroni corrected p-value of 0.0002.

| Preterm | | | | |
|---|---|---|---|---|
| **Marker** | **Direction** | **Significant** | **O.R [95% CI], p** | **Prevalence/PPV/NPV** |
| | Above range | 0 | 1.38 [1.06, 1.79] p=0.0167154 | 0.06 / 0.08 / 0.95 |
| | Two-sided | 0 | 1.13 [0.91, 1.4] p=0.2632332 | 0.06 / 0.06 / 0.95 |
| HCT | Below range | 0 | 0.83 [0.58, 1.2] p=0.326028 | 0.06 / 0.05 / 0.94 |
| | Above range | 1 | 1.78 [1.42, 2.25] p=8e-07 | 0.06 / 0.09 / 0.95 |
| | Two-sided | 1 | 1.48 [1.22, 1.79] p=6.47e-05 | 0.06 / 0.08 / 0.95 |
| HGB | Below range | 0 | 1.06 [0.76, 1.47] p=0.7298363 | 0.06 / 0.06 / 0.94 |
| | Above range | 1 | 1.98 [1.58, 2.48] p=2.5e-09 | 0.06 / 0.11 / 0.95 |
| | Two-sided | 1 | 1.51 [1.25, 1.82] p=1.76e-05 | 0.06 / 0.08 / 0.95 |
| RBC | Below range | 0 | 0.92 [0.66, 1.3] p=0.6498305 | 0.06 / 0.05 / 0.94 |
| | Above range | 0 | 1.17 [0.87, 1.56] p=0.2992647 | 0.06 / 0.07 / 0.95 |
| | Two-sided | 0 | 1.02 [0.81, 1.27] p=0.8762812 | 0.06 / 0.06 / 0.94 |
| WBC | Below range | 0 | 0.85 [0.6, 1.2] p=0.3569598 | 0.06 / 0.05 / 0.94 |
| | Above range | 0 | 1.06 [0.79, 1.41] p=0.714807 | 0.06 / 0.07 / 0.95 |
| | Two-sided | 0 | 1.07 [0.86, 1.32] p=0.5407211 | 0.06 / 0.06 / 0.95 |
| PLT | Below range | 0 | 1.08 [0.8, 1.46] p=0.6122308 | 0.06 / 0.06 / 0.94 |
| | Above range | 0 | 1.03 [0.72, 1.46] p=0.8757671 | 0.06 / 0.06 / 0.94 |
| | Two-sided | 0 | 1.18 [0.95, 1.47] p=0.1379957 | 0.06 / 0.07 / 0.95 |
| MCV | Below range | 0 | 1.29 [0.98, 1.7] p=0.0746969 | 0.06 / 0.07 / 0.95 |
| | Above range | 0 | 1.04 [0.75, 1.44] p=0.8023327 | 0.06 / 0.06 / 0.94 |
| | Two-sided | 0 | 1.16 [0.93, 1.44] p=0.191505 | 0.06 / 0.06 / 0.95 |
| MCH | Below range | 0 | 1.25 [0.93, 1.68] p=0.1310541 | 0.06 / 0.07 / 0.95 |

| | | | | |
|---|---|---|---|---|
| RDW | Above range | 0 | 1.14 [0.86, 1.53] p=0.3673808 | 0.06 / 0.07 / 0.95 |
| | Two-sided | 0 | 1.05 [0.83, 1.32] p=0.6756207 | 0.06 / 0.06 / 0.94 |
| | Below range | 0 | 0.92 [0.63, 1.34] p=0.6772089 | 0.06 / 0.05 / 0.94 |
| MCHC | Above range | 0 | 1.01 [0.74, 1.39] p=0.9434892 | 0.06 / 0.05 / 0.94 |
| | Two-sided | 0 | 0.99 [0.79, 1.24] p=0.9053749 | 0.06 / 0.06 / 0.94 |
| | Below range | 0 | 0.96 [0.7, 1.32] p=0.8228504 | 0.06 / 0.06 / 0.94 |

**Table S9. CBC index values are not associated with small for gestational age**. Pregnancies with a CBC at 26-29 weeks' gestation were considered (N=34,159). The odds ratios of developing examined complications are shown in table for a CBC index value above, outside, or below the study intervals at 26-29 weeks' gestation. Significant odds ratios are marked with a 1 in the corresponding column. Prevalence, positive predictive value (PPV), and negative predictive value (NPV) are also reported. Significance was evaluated with a Bonferroni corrected p-value of 0.0002.

| SGA | | | | |
|---|---|---|---|---|
| **Marker** | **Direction** | **Significant** | **O.R [95% CI], p** | **Prevalence/PPV/NPV** |
| HCT | Above range | 0 | 1.37 [1.08, 1.73] p=0.0099168 | 0.07 / 0.09 / 0.93 |
| | Two-sided | 0 | 1.17 [0.97, 1.41] p=0.101685 | 0.07 / 0.08 / 0.93 |
| | Below range | 0 | 0.95 [0.71, 1.27] p=0.7263295 | 0.07 / 0.07 / 0.93 |
| HGB | Above range | 0 | 1.38 [1.09, 1.74] p=0.0075311 | 0.07 / 0.09 / 0.93 |
| | Two-sided | 0 | 1.17 [0.97, 1.41] p=0.1076053 | 0.07 / 0.08 / 0.93 |
| | Below range | 0 | 0.92 [0.68, 1.23] p=0.5561933 | 0.07 / 0.07 / 0.93 |
| RBC | Above range | 0 | 1.4 [1.11, 1.78] p=0.0050601 | 0.07 / 0.1 / 0.93 |
| | Two-sided | 0 | 1.2 [0.99, 1.44] p=0.0568391 | 0.07 / 0.09 / 0.93 |
| | Below range | 0 | 0.97 [0.73, 1.29] p=0.8430721 | 0.07 / 0.08 / 0.93 |
| WBC | Above range | 0 | 1.33 [1.03, 1.71] p=0.027207 | 0.07 / 0.09 / 0.93 |
| | Two-sided | 0 | 1.25 [1.04, 1.51] p=0.0193946 | 0.07 / 0.09 / 0.93 |
| | Below range | 0 | 1.16 [0.88, 1.52] p=0.2953058 | 0.07 / 0.08 / 0.93 |
| PLT | Above range | 0 | 1.15 [0.89, 1.49] p=0.2758328 | 0.07 / 0.08 / 0.93 |
| | Two-sided | 0 | 1.14 [0.94, 1.38] p=0.1692594 | 0.07 / 0.08 / 0.93 |
| | Below range | 0 | 1.12 [0.85, 1.48] p=0.4101934 | 0.07 / 0.07 / 0.93 |
| MCV | Above range | 0 | 0.85 [0.61, 1.18] p=0.320099 | 0.07 / 0.07 / 0.93 |
| | Two-sided | 0 | 1.09 [0.89, 1.32] p=0.4104057 | 0.07 / 0.08 / 0.93 |
| | Below range | 0 | 1.28 [1.0, 1.64] p=0.0474822 | 0.07 / 0.09 / 0.93 |
| MCH | Above range | 0 | 0.87 [0.64, 1.18] p=0.37445 | 0.07 / 0.07 / 0.93 |
| | Two-sided | 0 | 1.01 [0.83, 1.23] p=0.9187892 | 0.07 / 0.08 / 0.93 |
| | Below range | 0 | 1.14 [0.87, 1.48] p=0.3428413 | 0.07 / 0.09 / 0.93 |
| RDW | Above range | 0 | 1.06 [0.81, 1.4] p=0.6673344 | 0.07 / 0.08 / 0.93 |
| | Two-sided | 0 | 1.12 [0.92, 1.37] p=0.2566016 | 0.07 / 0.09 / 0.93 |

| | | | | |
|---|---|---|---|---|
| | Below range | 0 | 1.18 [0.9, 1.57] p=0.2343965 | 0.07 / 0.1 / 0.93 |
| MCHC | Above range | 0 | 0.99 [0.75, 1.32] p=0.953062 | 0.07 / 0.06 / 0.93 |
| | Two-sided | 0 | 1.12 [0.92, 1.36] p=0.2444742 | 0.07 / 0.08 / 0.93 |
| | Below range | 0 | 1.23 [0.95, 1.6] p=0.1134315 | 0.07 / 0.09 / 0.93 |

**Table S10.Low hemoglobin is associated with transfusion**. Pregnancies with a CBC at 26-29 weeks' gestation were considered (N=34,159). The odds ratios of developing examined complications are shown in table for a CBC index value above, outside, or below the study intervals at 26-29 weeks' gestation. Significant odds ratios are marked with a 1 in the corresponding column. Prevalence, positive predictive value (PPV), and negative predictive value (NPV) are also reported. Significance was evaluated with a Bonferroni corrected p-value of 0.0002.

| **Transfusion** | | | | |
|---|---|---|---|---|
| **Marker** | **Direction** | **Significant** | **O.R [95% CI], p** | **Prevalence/PPV/NPV** |
| HCT | Above range | 0 | 0.49 [0.16, 1.54] p=0.2222441 | 0.01 / 0.0 / 0.99 |
| | Two-sided | 0 | 1.42 [0.84, 2.4] p=0.190891 | 0.01 / 0.01 / 0.99 |
| | Below range | 0 | 2.63 [1.46, 4.72] p=0.001271 | 0.01 / 0.02 / 0.99 |
| HGB | Above range | 0 | 0.64 [0.24, 1.7] p=0.3672356 | 0.01 / 0.0 / 0.99 |
| | Two-sided | 0 | 1.8 [1.13, 2.88] p=0.0138464 | 0.01 / 0.01 / 0.99 |
| | Below range | 1 | 3.39 [1.99, 5.76] p=6.4e-06 | 0.01 / 0.02 / 0.99 |
| RBC | Above range | 0 | 0.75 [0.31, 1.83] p=0.5309766 | 0.01 / 0.01 / 0.99 |
| | Two-sided | 0 | 1.64 [1.0, 2.69] p=0.0484014 | 0.01 / 0.01 / 0.99 |
| | Below range | 0 | 2.85 [1.6, 5.07] p=0.0003607 | 0.01 / 0.02 / 0.99 |
| WBC | Above range | 0 | 1.3 [0.6, 2.79] p=0.5049267 | 0.01 / 0.01 / 0.99 |
| | Two-sided | 0 | 1.9 [1.19, 3.02] p=0.0067911 | 0.01 / 0.01 / 0.99 |
| | Below range | 0 | 2.47 [1.37, 4.46] p=0.0026501 | 0.01 / 0.02 / 0.99 |
| PLT | Above range | 0 | 1.6 [0.84, 3.05] p=0.1499602 | 0.01 / 0.01 / 0.99 |
| | Two-sided | 0 | 1.56 [0.96, 2.54] p=0.0737547 | 0.01 / 0.01 / 0.99 |
| | Below range | 0 | 1.47 [0.72, 2.99] p=0.2922422 | 0.01 / 0.01 / 0.99 |
| MCV | Above range | 0 | 1.88 [0.92, 3.85] p=0.0832078 | 0.01 / 0.01 / 0.99 |
| | Two-sided | 0 | 1.75 [1.07, 2.87] p=0.0271086 | 0.01 / 0.01 / 0.99 |
| | Below range | 0 | 1.61 [0.83, 3.14] p=0.1614296 | 0.01 / 0.01 / 0.99 |
| MCH | Above range | 0 | 1.44 [0.67, 3.09] p=0.3518385 | 0.01 / 0.01 / 0.99 |
| | Two-sided | 0 | 2.01 [1.27, 3.18] p=0.0029315 | 0.01 / 0.01 / 0.99 |
| | Below range | 0 | 2.46 [1.39, 4.35] p=0.0019239 | 0.01 / 0.02 / 0.99 |

| | | | | |
|---|---|---|---|---|
| RDW | Above range | 0 | 2.37 [1.35, 4.16] p=0.0025793 | 0.01 / 0.02 / 0.99 |
| | Two-sided | 0 | 1.41 [0.82, 2.43] p=0.2108539 | 0.01 / 0.01 / 0.99 |
| | Below range | 0 | N/A | N/A |
| MCHC | Above range | 0 | 0.41 [0.1, 1.65] p=0.2091127 | 0.01 / 0.0 / 0.99 |
| | Two-sided | 0 | 1.22 [0.69, 2.14] p=0.4926457 | 0.01 / 0.01 / 0.99 |
| | Below range | 0 | 1.91 [1.03, 3.53] p=0.0391964 | 0.01 / 0.01 / 0.99 |

**Table S11. Elevated CBC index values are associated with HDP**. Pregnancies with a CBC at 26-29 weeks' gestation were considered (N=34,159), and those with abnormal tests for HDP or preeclampsia diagnosis before 29 weeks' gestation were excluded (N=254). The odds ratios of developing examined complications are shown in table for a CBC index value above, outside, or below the study intervals at 26-29 weeks' gestation. Significant odds ratios are marked with a 1 in the corresponding column. Prevalence, positive predictive value (PPV), and negative predictive value (NPV) are also reported. Significance was evaluated with a Bonferroni corrected p-value of 0.0002.

| HDP | | | | |
|---|---|---|---|---|
| **Marker** | **Direction** | **Significant** | **O.R [95% CI], p** | **Prevalence/PPV/NPV** |
| HCT | Above range | 1 | 1.67 [1.34, 2.09] p=6e-06 | 0.06 / 0.11 / 0.94 |
| | Two-sided | 0 | 1.38 [1.14, 1.66] p=0.0007458 | 0.06 / 0.08 / 0.94 |
| | Below range | 0 | 0.93 [0.67, 1.31] p=0.6874767 | 0.06 / 0.05 / 0.94 |
| HGB | Above range | 1 | 1.74 [1.4, 2.16] p=7e-07 | 0.06 / 0.11 / 0.94 |
| | Two-sided | 0 | 1.39 [1.15, 1.67] p=0.0006813 | 0.06 / 0.08 / 0.94 |
| | Below range | 0 | 0.83 [0.58, 1.2] p=0.3239622 | 0.06 / 0.04 / 0.94 |
| RBC | Above range | 1 | 1.74 [1.38, 2.18] p=1.8e-06 | 0.06 / 0.12 / 0.94 |
| | Two-sided | 0 | 1.26 [1.04, 1.53] p=0.0182904 | 0.06 / 0.08 / 0.94 |
| | Below range | 0 | 0.62 [0.42, 0.94] p=0.0236781 | 0.06 / 0.04 / 0.94 |
| WBC | Above range | 0 | 1.22 [0.95, 1.57] p=0.1150158 | 0.06 / 0.1 / 0.94 |
| | Two-sided | 0 | 1.06 [0.86, 1.3] p=0.5942563 | 0.06 / 0.07 / 0.94 |
| | Below range | 0 | 0.79 [0.55, 1.14] p=0.2092833 | 0.06 / 0.04 / 0.94 |
| PLT | Above range | 0 | 1.38 [1.09, 1.75] p=0.006591 | 0.06 / 0.1 / 0.94 |
| | Two-sided | 0 | 1.25 [1.04, 1.51] p=0.0169723 | 0.06 / 0.08 / 0.94 |
| | Below range | 0 | 1.05 [0.78, 1.42] p=0.7353264 | 0.06 / 0.05 / 0.94 |
| MCV | Above range | 0 | 0.67 [0.45, 1.01] p=0.0556196 | 0.06 / 0.04 / 0.94 |
| | Two-sided | 0 | 0.9 [0.72, 1.14] p=0.3972682 | 0.06 / 0.06 / 0.94 |
| | Below range | 0 | 1.08 [0.81, 1.44] p=0.584841 | 0.06 / 0.07 / 0.94 |
| MCH | Above range | 0 | 0.8 [0.57, 1.13] p=0.205486 | 0.06 / 0.05 / 0.94 |
| | Two-sided | 0 | 0.84 [0.66, 1.06] p=0.1430061 | 0.06 / 0.05 / 0.94 |
| | Below range | 0 | 0.87 [0.63, 1.21] p=0.4155704 | 0.06 / 0.06 / 0.94 |

| | | | | |
|---|---|---|---|---|
| RDW | Above range | 0 | 0.76 [0.55, 1.04] p=0.0850743 | 0.06 / 0.06 / 0.94 |
| | Two-sided | 0 | 0.86 [0.68, 1.08] p=0.1953094 | 0.06 / 0.06 / 0.94 |
| | Below range | 0 | 1.02 [0.72, 1.43] p=0.9177805 | 0.06 / 0.06 / 0.94 |
| MCHC | Above range | 0 | 1.1 [0.82, 1.47] p=0.5264104 | 0.06 / 0.06 / 0.94 |
| | Two-sided | 0 | 1.05 [0.86, 1.3] p=0.6296227 | 0.06 / 0.06 / 0.94 |
| | Below range | 0 | 1.01 [0.76, 1.35] p=0.9481458 | 0.06 / 0.07 / 0.94 |

**Table S12. Elevated CBC index values are associated with preeclampsia**. Pregnancies with a CBC at 26-29 weeks' gestation were considered (N=34,159), and those with abnormal tests for HDP or preeclampsia diagnosis before 29 weeks' gestation were excluded (N=254). The odds ratios of developing examined complications are shown in table for a CBC index value above, outside, or below the study intervals at 26-29 weeks' gestation. Significant odds ratios are marked with a 1 in the corresponding column. Prevalence, positive predictive value (PPV), and negative predictive value (NPV) are also reported. Significance was evaluated with a Bonferroni corrected p-value of 0.0002.

| **Preeclampsia** | | | | |
|---|---|---|---|---|
| **Marker** | **Direction** | **Significant** | **O.R [95% CI], p** | **Prevalence/PPV/NPV** |
| HCT | Above range | 0 | 1.68 [1.23, 2.3] p=0.0010495 | 0.03 / 0.05 / 0.97 |
| | Two-sided | 0 | 1.41 [1.09, 1.83] p=0.0094056 | 0.03 / 0.04 / 0.97 |
| | Below range | 0 | 1.0 [0.64, 1.58] p=0.9956481 | 0.03 / 0.03 / 0.97 |
| HGB | Above range | 1 | 1.91 [1.43, 2.54] p=1.18e-05 | 0.03 / 0.06 / 0.97 |
| | Two-sided | 1 | 1.63 [1.28, 2.08] p=8.34e-05 | 0.03 / 0.05 / 0.97 |
| | Below range | 0 | 1.17 [0.76, 1.8] p=0.4785205 | 0.03 / 0.03 / 0.97 |
| RBC | Above range | 1 | 2.13 [1.62, 2.82] p=1e-07 | 0.03 / 0.07 / 0.97 |
| | Two-sided | 1 | 1.69 [1.33, 2.14] p=1.56e-05 | 0.03 / 0.05 / 0.97 |
| | Below range | 0 | 0.97 [0.61, 1.55] p=0.9046784 | 0.03 / 0.03 / 0.97 |
| WBC | Above range | 0 | 1.22 [0.87, 1.7] p=0.2519176 | 0.03 / 0.05 / 0.97 |
| | Two-sided | 0 | 1.07 [0.81, 1.41] p=0.6549407 | 0.03 / 0.03 / 0.97 |
| | Below range | 0 | 0.82 [0.5, 1.36] p=0.4500363 | 0.03 / 0.02 / 0.97 |
| PLT | Above range | 0 | 1.6 [1.19, 2.16] p=0.001943 | 0.03 / 0.06 / 0.97 |
| | Two-sided | 1 | 1.59 [1.26, 2.01] p=8.42e-05 | 0.03 / 0.05 / 0.97 |
| | Below range | 0 | 1.53 [1.06, 2.2] p=0.0220363 | 0.03 / 0.04 / 0.97 |
| MCV | Above range | 0 | 0.67 [0.4, 1.12] p=0.1278023 | 0.03 / 0.02 / 0.97 |
| | Two-sided | 0 | 1.09 [0.81, 1.47] p=0.5607385 | 0.03 / 0.04 / 0.97 |
| | Below range | 0 | 1.36 [0.96, 1.92] p=0.0817266 | 0.03 / 0.05 / 0.97 |
| MCH | Above range | 0 | 0.99 [0.63, 1.56] p=0.9656853 | 0.03 / 0.03 / 0.97 |
| | Two-sided | 0 | 1.06 [0.79, 1.43] p=0.6908829 | 0.03 / 0.03 / 0.97 |
| | Below range | 0 | 1.11 [0.75, 1.65] p=0.5893602 | 0.03 / 0.04 / 0.97 |

| | | | | |
|---|---|---|---|---|
| RDW | Above range | 0 | 0.86 [0.57, 1.28] p=0.4496105 | 0.03 / 0.04 / 0.97 |
| | Two-sided | 0 | 0.91 [0.66, 1.24] p=0.5349388 | 0.03 / 0.03 / 0.97 |
| | Below range | 0 | 0.99 [0.61, 1.63] p=0.9788633 | 0.03 / 0.03 / 0.97 |
| MCHC | Above range | 0 | 1.05 [0.7, 1.58] p=0.818664 | 0.03 / 0.03 / 0.97 |
| | Two-sided | 0 | 1.16 [0.87, 1.53] p=0.3114627 | 0.03 / 0.03 / 0.97 |
| | Below range | 0 | 1.24 [0.86, 1.79] p=0.2467939 | 0.03 / 0.04 / 0.97 |

**S13. Odds ratios and PPVs for extreme CBC index values are consistent in an out-of-sample validation cohort.** We analyzed associations between adverse outcomes and the presence of a CBC index above, outside, or below the study reference intervals derived in discovery at 26-29 weeks' gestation. Only combinations of CBC index/adverse outcomes that were significant in the discovery cohort were considered for this analysis. In the table we compare validation and discovery cohort results in terms of ORs, prevalence, positive predictive value (PPV), and negative predictive value (NPV). Significance was evaluated with a Bonferroni corrected p-value of 0.0002, and significant ORs in validation are marked with a 1 in the corresponding column.

| Marker | Direction | Validation Significant? | Validation OR | Discovery OR | Validation Prevalence/PPV/NPV | Discovery Prevalence/PPV/NPV |
|---|---|---|---|---|---|---|
| Composite | | | | | | |
| HCT | Above range | 1 | 1.33 [1.21, 1.46] p=0.0 | 1.42 [1.21, 1.67] p=1.8e-05 | 0.22 / 0.28 / 0.78 | 0.17 / 0.23 / 0.83 |
| HGB | Above range | 1 | 1.53 [1.32, 1.76] p=0.0 | 1.65 [1.42, 1.92] p=1.4e-10 | 0.22 / 0.3 / 0.78 | 0.17 / 0.26 / 0.83 |
| HGB | Two-sided | 1 | 1.18 [1.09, 1.29] p=0.0001207 | 1.34 [1.18, 1.52] p=4.4e-06 | 0.22 / 0.25 / 0.78 | 0.17 / 0.22 / 0.83 |
| RBC | Above range | 1 | 1.53 [1.37, 1.7] p=0.0 | 1.61 [1.37, 1.88] p=3.9e-09 | 0.22 / 0.33 / 0.78 | 0.17 / 0.26 / 0.83 |
| HDP | | | | | | |
| HCT | Above range | 1 | 1.46 [1.31, 1.64] p=0.0 | 1.67 [1.34, 2.09] p=6e-06 | 0.12 / 0.18 / 0.88 | 0.06 / 0.11 / 0.94 |
| HGB | Above range | 1 | 1.61 [1.36, 1.9] p=0.0 | 1.74 [1.4, 2.16] p=7e-07 | 0.12 / 0.19 / 0.88 | 0.06 / 0.11 / 0.94 |
| RBC | Above range | 1 | 1.53 [1.35, 1.75] p=0.0 | 1.74 [1.38, 2.18] p=1.8e-06 | 0.12 / 0.2 / 0.88 | 0.06 / 0.12 / 0.94 |
| Preterm | | | | | | |
| HGB | Above range | 0 | 1.5 [1.18, 1.91] p=0.0008903 | 1.78 [1.42, 2.25] p=8e-07 | 0.06 / 0.08 / 0.94 | 0.06 / 0.09 / 0.95 |
| HGB | Two-sided | 0 | 1.3 [1.13, 1.49] p=0.0003137 | 1.48 [1.22, 1.79] p=6.47e-05 | 0.06 / 0.07 / 0.94 | 0.06 / 0.08 / 0.95 |
| RBC | Above range | 1 | 1.51 [1.27, 1.8] p=3.7e-06 | 1.98 [1.58, 2.48] p=2.5e-09 | 0.06 / 0.09 / 0.94 | 0.06 / 0.11 / 0.95 |

| | | | | | | |
|---|---|---|---|---|---|---|
| RBC | Two-sided | 1 | 1.36 [1.18, 1.56] p=1.53e-05 | 1.51 [1.25, 1.82] p=1.76e-05 | 0.06 / 0.08 / 0.94 | 0.06 / 0.08 / 0.95 |

**Table S14. Pregnancies with rare longitudinal changes between 7-14 and 26-29 weeks' gestation are at higher risk of complications.** Rare change thresholds are reported in the following units: HGB (g/dL), HCT (%), RBC ($10^6$/mm$^3$), PLT ($10^3$/μL), MCV (fL), WBC ($10^3$/μL), RDW (%), MCH (pg), MCHC (g/dL). Significance was evaluated with a Bonferroni corrected p-value of $p < 9 \times 10^{-6}$. In the table we report the odds ratio, the number of pregnancies with the rare behavior (Total N), the number of complicated pregnancies with a rare behavior (Complicated N), the number of complicated pregnancies with a rare behavior that fall within the reference interval at 26-29 weeks' gestation (Complicated N in range), and their percentage and associated confidence interval. We also report the prevalence of the condition in the dataset, the positive predictive value (PPV), and the negative predictive value (NPV).

| Marker | Direction | Threshold | OR [95% CI] | Total N | Complicated N | Complicate N in range (N [%]) | Complicated N in range (% [95% CI]) | Prevalence [95% CI] | Recall [95% CI] | 2-=llllllll | NPV [95% CI] |
|---|---|---|---|---|---|---|---|---|---|---|---|
| **Composite** | | | | | | | | | | | |
| HCT | Increase | 1.801 | 1.59 [1.31, 1.94], p=4e-06 | 519 | 129 | 101 (78) | 78 [71, 85] | 16.8 [16.4, 17.2] | 2.6 [2.2, 3.1] | 24.9 [21.2, 28.8] | 83.3 [82.9, 83.7] |
| HGB | Increase | 0.667 | 1.99 [1.57, 2.52], p=1.42e-08 | 328 | 98 | 80 (82) | 82 [73, 89] | 16.8 [16.4, 17.2] | 2.0 [1.6, 2.4] | 29.9 [25.1, 34.8] | 83.3 [82.9, 83.7] |
| WBC | Decrease | N/A | N/A | N/A | N/A | N/A | N/A | N/A | N/A | N/A | N/A |
| RBC | Increase | 0.071 | 1.94 [1.6, 2.35], p=2.19e-11 | 526 | 152 | 124 (82) | 82 [76, 87] | 16.8 [16.4, 17.3] | 3.1 [2.6, 3.6] | 28.9 [25.0, 32.9] | 83.4 [82.9, 83.8] |
| PLT | Increase | -0.051 | 1.2 [1.12, 1.29], p=2.57e-07 | 6970 | 1359 | 1238 (91) | 91 [90, 93] | 16.8 [16.4, 17.3] | 27.7 [26.4, 29.0] | 19.5 [18.5, 20.4] | 84.0 [83.5, 84.5] |
| MCV | Decrease | -0.747 | 1.32 [1.2, 1.45], p=3.51e-09 | 3349 | 704 | 656 (93) | 93 [91, 95] | 16.8 [16.4, 17.2] | 14.3 [13.4, 15.4] | 21.0 [19.7, 22.4] | 83.7 [83.3, 84.2] |
| MCH | Decrease | N/A | N/A | N/A | N/A | N/A | N/A | N/A | N/A | N/A | N/A |

| Marker | Direction | Threshold | OR [95% CI] | Total N | Complicated N | Complicate N in range (N [%]) | Complicated N in range (% [95% CI]) | Prevalence [95% CI] | Recall [95% CI] | PPV [95% CI] | NPV [95% CI] |
|---|---|---|---|---|---|---|---|---|---|---|---|
| RDW | Decrease | -2.209 | 1.42 [1.05, 1.92], p=0.022981 | 237 | 56 | 49 (88) | 88 [78, 96] | 16.8 [16.4, 17.2] | 1.1 [0.9, 1.4] | 23.6 [18.4, 28.8] | 83.2 [82.8, 83.6] |
| MCHC | Increase | 3.791 | 2.9 [0.65, 13.01], p=0.163537 | 7 | 3 | 2 (67) | 67 [0, 100] | 16.8 [16.4, 17.2] | 0.1 [0.0, 0.1] | 42.9 [0.0, 83.3] | 83.2 [82.8, 83.6] |
| **Marker** | **Direction** | **Threshold** | **OR [95% CI]** | **Total N** | **Complicated N** | **Complicate N in range (N [%])** | **Complicated N in range (% [95% CI])** | **Prevalence [95% CI]** | **Recall [95% CI]** | **PPV [95% CI]** | **NPV [95% CI]** |
| **Preeclampsia** | | | | | | | | | | | |
| HCT | Increase | 5.498 | 2.69 [0.55, 13.09], p=0.221046 | 20 | 2 | 1 (50) | 50 [0, 100] | 3.1 [2.9, 3.3] | 0.2 [0.0, 0.6] | 10.0 [0.0, 26.1] | 96.9 [96.7, 97.1] |
| HGB | Increase | 3 | 6.13 [0.71, 53.22], p=0.099964 | 5 | 1 | 1 (100) | 100 [100, 100] | 3.1 [2.9, 3.3] | 0.1 [0.0, 0.3] | 20.0 [0.0, 66.7] | 96.9 [96.7, 97.1] |
| WBC | Decrease | -9.226 | 18.3 [2.13, 157.31], p=0.008079 | 4 | 1 | 1 (100) | 100 [100, 100] | 3.1 [2.9, 3.3] | 0.1 [0.0, 0.4] | 25.0 [0.0, 100.0] | 96.9 [96.7, 97.1] |
| RBC | Increase | -0.068 | 1.71 [1.36, 2.14], p=4e-06 | 1822 | 91 | 72 (79) | 79 [71, 88] | 3.1 [2.9, 3.3] | 10.1 [8.3, 12.1] | 5.0 [4.1, 6.1] | 97.1 [96.9, 97.2] |
| PLT | Increase | 170.939 | 7.54 [1.03, 55.5], p=0.047229 | 6 | 1 | 0 (0) | 0 [0, 0] | 3.1 [2.9, 3.3] | 0.1 [0.0, 0.4] | 16.7 [0.0, 50.0] | 96.9 [96.7, 97.1] |
| MCV | Decrease | 0.081 | 1.44 [1.24, 1.68], p=3e-06 | 6380 | 286 | 271 (95) | 95 [92, 97] | 3.1 [2.9, 3.3] | 31.9 [28.8, 35.1] | 4.5 [4.0, 5.0] | 97.3 [97.1, 97.5] |
| MCH | Decrease | N/A | N/A | N/A | N/A | N/A | N/A | N/A | N/A | N/A | N/A |
| RDW | Decrease | -4.391 | 4.27 [1.62, 11.26], p=0.00331 | 48 | 6 | 5 (83) | 83 [40, 100] | 3.1 [2.9, 3.3] | 0.7 [0.2, 1.2] | 12.5 [4.2, 22.2] | 96.9 [96.8, 97.1] |
| MCHC | Increase | 3.791 | 4.82 [0.64, 36.43], p=0.127679 | 7 | 1 | 1 (100) | 100 [100, 100] | 3.1 [2.9, 3.3] | 0.1 [0.0, 0.4] | 14.3 [0.0, 44.9] | 96.9 [96.7, 97.1] |
| **Marker** | **Direction** | **Threshold** | **OR [95% CI]** | **Total N** | **Complicated N** | **Complicate N in range (N [%])** | **Complicated N in range** | **Prevalence [95% CI]** | **Recall [95% CI]** | **PPV [95% CI]** | **NPV [95% CI]** |

| | | | | | | | | | | | |
|---|---|---|---|---|---|---|---|---|---|---|---|
| | | | | | | | **(% [95% CI])** | | | | |
| **HDP** | | | | | | | | | | | |
| HCT | Increase | -0.047 | 1.4 [1.21, 1.62], p=6e-06 | 2747 | 217 | 182 (84) | 84 [79, 89] | 6.4 [6.1, 6.7] | 11.7 [10.2, 13.3] | 7.9 [6.9, 8.9] | 93.8 [93.5, 94.1] |
| HGB | Increase | 0.111 | 1.66 [1.36, 2.02], p=1e-06 | 1310 | 122 | 100 (82) | 82 [74, 89] | 6.4 [6.1, 6.7] | 6.6 [5.5, 7.7] | 9.3 [7.7, 10.9] | 93.8 [93.5, 94.0] |
| WBC | Decrease | -9.226 | 10.55 [1.3, 85.9], p=0.02763 | 4 | 1 | 1 (100) | 100 [100, 100] | 6.4 [6.1, 6.7] | 0.1 [0.0, 0.2] | 25.0 [0.0, 100.0] | 93.6 [93.3, 93.9] |
| RBC | Increase | -0.034 | 1.54 [1.28, 1.85], p=5e-06 | 1471 | 139 | 110 (79) | 79 [72, 86] | 6.4 [6.1, 6.7] | 7.5 [6.3, 8.7] | 9.4 [8.0, 11.0] | 93.8 [93.5, 94.1] |
| PLT | Increase | N/A | N/A | N/A | N/A | N/A | N/A | N/A | N/A | N/A | N/A |
| MCV | Decrease | -0.747 | 1.35 [1.18, 1.55], p=9e-06 | 3349 | 313 | 292 (93) | 93 [90, 96] | 6.4 [6.1, 6.7] | 16.8 [15.2, 18.5] | 9.3 [8.4, 10.3] | 94.0 [93.7, 94.3] |
| MCH | Decrease | N/A | N/A | N/A | N/A | N/A | N/A | N/A | N/A | N/A | N/A |
| RDW | Decrease | -7.118 | 5.2 [1.1, 24.48], p=0.037154 | 9 | 2 | 2 (100) | 100 [100, 100] | 6.4 [6.1, 6.7] | 0.1 [0.0, 0.3] | 22.2 [0.0, 53.9] | 93.6 [93.3, 93.9] |
| MCHC | Increase | 3.791 | 7.74 [1.39, 43.0], p=0.019371 | 7 | 2 | 1 (50) | 50 [0, 100] | 6.4 [6.1, 6.7] | 0.1 [0.0, 0.3] | 28.6 [0.0, 66.8] | 93.6 [93.3, 93.9] |
| **Marker** | **Direction** | **Threshold** | **OR [95% CI]** | **Total N** | **Complicated N** | **Complicate N in range (N [%])** | **Complicated N in range (% [95% CI])** | **Prevalence [95% CI]** | **Recall [95% CI]** | **PPV [95% CI]** | **NPV [95% CI]** |
| **Preterm** | | | | | | | | | | | |
| HCT | Increase | 0.261 | 1.47 [1.24, 1.74], p=9e-06 | 2153 | 162 | 136 (84) | 84 [78, 90] | 5.4 [5.2, 5.7] | 10.2 [8.7, 11.6] | 7.5 [6.4, 8.7] | 94.7 [94.5, 95.0] |

| | | | | | | | | | | | |
|---|---|---|---|---|---|---|---|---|---|---|---|
| HGB | Increase | 0.667 | 2.28 [1.62, 3.2], p=2e-06 | 328 | 39 | 30 (77) | 77 [64, 90] | 5.4 [5.2, 5.7] | 2.5 [1.7, 3.2] | 11.9 [8.3, 15.3] | 94.6 [94.4, 94.9] |
| WBC | Decrease | -9.226 | 7.01 [0.76, 65.04], p=0.0867 | 4 | 1 | 1 (100) | 100 [100, 100] | 5.4 [5.2, 5.7] | 0.1 [0.0, 0.2] | 25.0 [0.0, 100.0] | 94.6 [94.3, 94.8] |
| RBC | Increase | 0.071 | 2.33 [1.77, 3.07], p=1.41e-09 | 526 | 62 | 50 (81) | 81 [71, 90] | 5.4 [5.2, 5.7] | 3.9 [2.9, 4.9] | 11.8 [8.9, 14.5] | 94.7 [94.4, 94.9] |
| PLT | Increase | 112.162 | 3.58 [1.58, 8.12], p=0.002307 | 44 | 8 | 4 (50) | 50 [14, 89] | 5.4 [5.2, 5.7] | 0.5 [0.2, 0.9] | 18.2 [7.1, 30.2] | 94.6 [94.3, 94.8] |
| MCV | Decrease | -0.747 | 1.41 [1.22, 1.63], p=3e-06 | 3349 | 251 | 231 (92) | 92 [89, 95] | 5.4 [5.2, 5.7] | 15.8 [14.1, 17.6] | 7.5 [6.6, 8.3] | 94.8 [94.6, 95.1] |
| MCH | Decrease | -3.685 | 2.65 [0.64, 11.08], p=0.180649 | 18 | 2 | 0 (0) | 0 [0, 0] | 5.4 [5.2, 5.7] | 0.1 [0.0, 0.3] | 11.1 [0.0, 28.0] | 94.6 [94.3, 94.8] |
| RDW | Decrease | -1.118 | 1.2 [0.91, 1.59], p=0.205246 | 850 | 56 | 49 (88) | 88 [78, 95] | 5.4 [5.2, 5.7] | 3.5 [2.7, 4.4] | 6.6 [5.0, 8.2] | 94.6 [94.3, 94.9] |
| MCHC | Increase | 3.427 | 2.91 [0.65, 13.04], p=0.163782 | 13 | 2 | 2 (100) | 100 [100, 100] | 5.4 [5.2, 5.7] | 0.1 [0.0, 0.3] | 15.4 [0.0, 37.5] | 94.6 [94.3, 94.8] |
| **Marker** | **Direction** | **Threshold** | **OR [95% CI]** | **Total N** | **Complicated N** | **Complicate N in range (N [%])** | **Complicated N in range (% [95% CI])** | **Prevalence [95% CI]** | **Recall [95% CI]** | **PPV [95% CI]** | **NPV [95% CI]** |
| **SGA** | | | | | | | | | | | |
| HCT | Increase | 1.185 | 1.67 [1.35, 2.05], p=2e-06 | 986 | 116 | 93 (80) | 80 [73, 87] | 6.8 [6.5, 7.1] | 5.9 [4.8, 7.0] | 11.8 [9.9, 13.9] | 93.4 [93.1, 93.7] |
| HGB | Increase | 0.222 | 1.64 [1.34, 2.01], p=2e-06 | 1004 | 119 | 104 (87) | 87 [81, 93] | 6.8 [6.5, 7.1] | 6.0 [5.0, 7.1] | 11.9 [9.9, 14.0] | 93.4 [93.1, 93.7] |
| WBC | Decrease | N/A | N/A | N/A | N/A | N/A | N/A | N/A | N/A | N/A | N/A |
| RBC | Increase | 0.071 | 1.92 [1.47, 2.51], p=2e-06 | 526 | 69 | 60 (87) | 87 [78, 94] | 6.8 [6.5, 7.1] | 3.5 [2.7, 4.3] | 13.1 [10.2, 16.2] | 93.3 [93.0, 93.6] |

| | | | | | | | | | | | |
|---|---|---|---|---|---|---|---|---|---|---|---|
| PLT | Increase | 122.848 | 2.16 [0.88, 5.28], p=0.092028 | 33 | 5 | 2 (40) | 40 [0, 100] | 6.8 [6.5, 7.1] | 0.3 [0.1, 0.5] | 15.2 [3.4, 29.0] | 93.2 [92.9, 93.5] |
| MCV | Decrease | 0.909 | 1.31 [1.17, 1.45], p=2e-06 | 6725 | 508 | 488 (96) | 96 [94, 98] | 6.8 [6.5, 7.1] | 25.7 [23.9, 27.6] | 7.6 [7.0, 8.2] | 93.4 [93.1, 93.8] |
| MCH | Decrease | N/A | N/A | N/A | N/A | N/A | N/A | N/A | N/A | N/A | N/A |
| RDW | Decrease | -4.209 | 2.03 [0.89, 4.62], p=0.091902 | 51 | 8 | 6 (75) | 75 [40, 100] | 6.8 [6.5, 7.1] | 0.4 [0.2, 0.7] | 15.7 [6.8, 26.8] | 93.2 [92.9, 93.5] |
| MCHC | Increase | -0.209 | 1.23 [1.12, 1.35], p=8e-06 | 14127 | 1049 | 1007 (96) | 96 [95, 97] | 6.8 [6.5, 7.1] | 53.0 [50.9, 55.2] | 7.4 [7.0, 7.9] | 93.8 [93.4, 94.2] |
| **Marker** | **Direction** | **Threshold** | **OR [95% CI]** | **Total N** | **Complicated N** | **Complicate N in range (N [%])** | **Complicated N in range (% [95% CI])** | **Prevalence [95% CI]** | **Recall [95% CI]** | **PPV [95% CI]** | **NPV [95% CI]** |
| **Transfusion** | | | | | | | | | | | |
| HCT | Increase | 2.417 | 1.52 [0.48, 4.78], p=0.47652 | 297 | 3 | 2 (67) | 67 [0, 100] | 0.7 [0.6, 0.8] | 1.5 [0.0, 3.4] | 1.0 [0.0, 2.3] | 99.3 [99.2, 99.4] |
| HGB | Increase | 0.444 | 1.32 [0.54, 3.23], p=0.542667 | 584 | 5 | 4 (80) | 80 [39, 100] | 0.7 [0.6, 0.8] | 2.5 [0.5, 4.9] | 0.9 [0.2, 1.7] | 99.3 [99.2, 99.4] |
| WBC | Decrease | -6.014 | 3.22 [0.44, 23.61], p=0.24947 | 46 | 1 | 1 (100) | 100 [100, 100] | 0.7 [0.6, 0.8] | 0.5 [0.0, 1.7] | 2.2 [0.0, 7.8] | 99.3 [99.2, 99.4] |
| RBC | Increase | N/A | N/A | N/A | N/A | N/A | N/A | N/A | N/A | N/A | N/A |
| PLT | Increase | N/A | N/A | N/A | N/A | N/A | N/A | N/A | N/A | N/A | N/A |
| MCV | Decrease | -6.96 | 3.69 [0.48, 28.29], p=0.209343 | 40 | 1 | 0 (0) | 0 [0, 0] | 0.7 [0.6, 0.8] | 0.5 [0.0, 1.6] | 2.5 [0.0, 8.8] | 99.3 [99.2, 99.4] |
| MCH | Decrease | -3.484 | 13.74 [3.08, 61.2], p=0.000588 | 24 | 2 | 0 (0) | 0 [0, 0] | 0.7 [0.6, 0.8] | 1.0 [0.0, 2.7] | 8.3 [0.0, 20.8] | 99.3 [99.2, 99.4] |

| | | | | | | | | | | | |
|---|---|---|---|---|---|---|---|---|---|---|---|
| RDW | Decrease | -4.573 | 3.53 [0.48, 25.94], p=0.215299 | 41 | 1 | 1 (100) | 100 [100, 100] | 0.7 [0.6, 0.8] | 0.5 [0.0, 1.7] | 2.4 [0.0, 7.9] | 99.3 [99.2, 99.4] |
| MCHC | Increase | 3.306 | 7.83 [1.06, 57.98], p=0.043876 | 18 | 1 | 0 (0) | 0 [0, 0] | 0.7 [0.6, 0.8] | 0.5 [0.0, 1.7] | 5.6 [0.0, 18.8] | 99.3 [99.2, 99.4] |

**Table S15. Odds ratios and PPVs for rare behaviors are consistent in an out-of-sample validation cohort in composite outcome.** Out of sample validation of rare dynamics for hemoglobin HGB (g/dL), red blood cell count RBC ($10^6$/mm$^3$), hematocrit HCT (%), platelet count PLT ($10^3$/µL), and mean red cell volume MCV (fL) shows odds ratios, PPV, and NPV consistent with the discovery cohort. The table reports the total number of pregnancies with rare longitudinal change between 7-14 and 26-29 weeks' gestation (Total N), the total number of pregnancies with the complication of interest, the rare longitudinal change (Complicated N), and the percentage of pregnancies with both the complication and a marker value within the reference interval for 26-29 weeks' gestation with associated confidence intervals (Complicated N in range %). These values are shown for both the discovery and validation cohorts. We also show the prevalence of each adverse outcome and the associated confidence interval obtained via bootstrapping, for both validation and discovery cohorts. Threshold for statistical significance of the odds ratio was $p<9\times10^{-6}$ with Bonferroni correction.

| **Marker** | HCT | HGB | RBC | PLT | MCV |
|---|---|---|---|---|---|
| **Direction** | Increase | Increase | Increase | Increase | Decrease |
| **Threshold** | 1.801 | 0.667 | 0.071 | -0.051 | -0.747 |
| **Validation OR** | 1.49 [1.3, 1.7], p=3.743652e-09 | 1.58 [1.33, 1.86], p=8.8089663e-08 | 1.69 [1.48, 1.93], p=1.4e-14 | 1.14 [1.08, 1.2], p=2.463420877e-06 | 1.34 [1.24, 1.45], p=5.7e-14 |
| **Discovery OR** | 1.59 [1.31, 1.94], p=4e-06 | 1.99 [1.57, 2.52], p=1.42e-08 | 1.94 [1.6, 2.35], p=2.19e-11 | 1.2 [1.12, 1.29], p=2.57e-07 | 1.32 [1.2, 1.45], p=3.51e-09 |
| **Discovery Total N** | 519 | 328 | 526 | 6970 | 3349 |
| **Validation Total N** | 1130 | 672 | 1033 | 10169 | 3786 |
| **Discovery Complicated N** | 129 | 98 | 152 | 1359 | 704 |
| **Validation Complicated N** | 334 | 210 | 334 | 2501 | 1086 |
| **Discovery Complicated N in range %** | 78 [71, 85] | 82 [73, 89] | 82 [76, 87] | 91 [90, 93] | 93 [91, 95] |
| **Validation Complicated N in range %** | 71 [66, 75] | 80 [75, 86] | 77 [72, 81] | 93 [93, 94] | 90 [88, 92] |
| **Discovery Prevalence** | 16.8 [16.4, 17.2] | 16.8 [16.4, 17.2] | 16.8 [16.4, 17.3] | 16.8 [16.4, 17.3] | 16.8 [16.4, 17.2] |
| **Validation Prevalence** | 22.1 [21.7, 22.5] | 22.1 [21.8, 22.5] | 22.1 [21.7, 22.4] | 22.1 [21.8, 22.5] | 22.1 [21.7, 22.5] |
| **Discovery PPV** | 24.9 [21.2, 28.8] | 29.9 [25.1, 34.8] | 28.9 [25.0, 32.9] | 19.5 [18.5, 20.4] | 21.0 [19.7, 22.4] |
| **Validation PPV** | 29.6 [26.9, 32.3] | 31.2 [27.9, 34.5] | 32.3 [29.6, 35.3] | 24.6 [23.8, 25.4] | 28.7 [27.2, 30.1] |
| **Discovery Recall** | 2.6 [2.2, 3.1] | 2.0 [1.6, 2.4] | 3.1 [2.6, 3.6] | 27.7 [26.4, 29.0] | 14.3 [13.4, 15.4] |
| **Validation Recall** | 3.0 [2.7, 3.3] | 1.9 [1.6, 2.1] | 3.0 [2.7, 3.3] | 22.4 [21.6, 23.1] | 9.7 [9.2, 10.3] |
| **Discovery NPV** | 83.3 [82.9, 83.7] | 83.3 [82.9, 83.7] | 83.4 [82.9, 83.8] | 84.0 [83.5, 84.5] | 83.7 [83.3, 84.2] |

| | | | | | |
|---|---|---|---|---|---|
| **Validation NPV** | 78.1 [77.7, 78.5] | 78.0 [77.7, 78.4] | 78.1 [77.8, 78.5] | 78.5 [78.1, 78.9] | 78.5 [78.1, 78.8] |

**Table S16. Odds ratios and PPVs for rare behaviors are consistent in an out-of-sample validation cohort in HDP.** Out of sample validation of rare dynamics for hemoglobin HGB (g/dL), red blood cell count RBC ($10^6$/mm$^3$), hematocrit HCT (%), platelet count PLT ($10^3$/μL), and mean red cell volume MCV (fL) shows odds ratios, PPV, and NPV consistent with the discovery cohort. The table reports the total number of pregnancies with rare longitudinal change between 7-14 and 26-29 weeks' gestation (Total N), the total number of pregnancies with the complication of interest, the rare longitudinal change (Complicated N), and the percentage of pregnancies with both the complication and a marker value within the reference interval for 26-29 weeks' gestation with associated confidence intervals (Complicated N in range %). These values are shown for both the discovery and validation cohorts. We also show the prevalence of each adverse outcome and the associated confidence interval obtained via bootstrapping, for both validation and discovery cohorts. Threshold for statistical significance of the odds ratio was $p<9 \times 10^{-6}$ with Bonferroni correction.

| **Marker** | HCT | HGB | RBC | MCV |
|---|---|---|---|---|
| **Direction** | Increase | Increase | Increase | Decrease |
| **Threshold** | -0.047 | 0.111 | -0.034 | -0.747 |
| **Validation OR** | 1.18 [1.08, 1.29], p=0.000243932839498 | 1.26 [1.11, 1.42], p=0.000229330049059 | 1.41 [1.26, 1.58], p=2.427698e-09 | 1.31 [1.2, 1.44], p=1.4052082e-08 |
| **Discovery OR** | 1.4 [1.21, 1.62], p=6e-06 | 1.66 [1.36, 2.02], p=1e-06 | 1.54 [1.28, 1.85], p=5e-06 | 1.35 [1.18, 1.55], p=9e-06 |
| **Discovery Total N** | 2747 | 1310 | 1471 | 3349 |
| **Validation Total N** | 5151 | 2425 | 2483 | 3786 |
| **Discovery Complicated N** | 217 | 122 | 139 | 313 |
| **Validation Complicated N** | 669 | 333 | 392 | 651 |
| **Discovery Complicated N in range %** | 84 [79, 89] | 82 [74, 89] | 79 [72, 86] | 93 [90, 96] |
| **Validation Complicated N in range %** | 78 [75, 81] | 83 [79, 87] | 80 [76, 83] | 92 [90, 94] |
| **Discovery Prevalence** | 6.4 [6.1, 6.7] | 6.4 [6.1, 6.7] | 6.4 [6.1, 6.7] | 6.4 [6.1, 6.7] |
| **Validation Prevalence** | 12.1 [11.8, 12.4] | 12.1 [11.8, 12.4] | 12.1 [11.8, 12.4] | 12.1 [11.8, 12.4] |
| **Discovery PPV** | 7.9 [6.9, 8.9] | 9.3 [7.7, 10.9] | 9.4 [8.0, 11.0] | 9.3 [8.4, 10.3] |
| **Validation PPV** | 13.0 [12.2, 13.9] | 13.7 [12.4, 15.0] | 15.8 [14.4, 17.3] | 17.2 [16.0, 18.4] |
| **Discovery Recall** | 11.7 [10.2, 13.3] | 6.6 [5.5, 7.7] | 7.5 [6.3, 8.7] | 16.8 [15.2, 18.5] |
| **Validation Recall** | 10.9 [10.2, 11.7] | 5.4 [4.9, 6.0] | 6.4 [5.8, 7.0] | 10.6 [9.9, 11.4] |
| **Discovery NPV** | 93.8 [93.5, 94.1] | 93.8 [93.5, 94.0] | 93.8 [93.5, 94.1] | 94.0 [93.7, 94.3] |

| Validation NPV | 88.0 [87.7, 88.3] | 88.0 [87.7, 88.3] | 88.1 [87.8, 88.4] | 88.3 [88.0, 88.6] |
|---|---|---|---|---|

**Table S17. Odds ratios and PPVs for rare behaviors are consistent in an out-of-sample validation cohort for preterm.** Out of sample validation of rare dynamics for hemoglobin HGB (g/dL), red blood cell count RBC ($10^6$/mm$^3$), hematocrit HCT (%), platelet count PLT ($10^3$/µL), and mean red cell volume MCV (fL) shows odds ratios, PPV, and NPV consistent with the discovery cohort. The table reports the total number of pregnancies with rare longitudinal change between 7-14 and 26-29 weeks' gestation (Total N), the total number of pregnancies with the complication of interest, the rare longitudinal change (Complicated N), and the percentage of pregnancies with both the complication and a marker value within the reference interval for 26-29 weeks' gestation with associated confidence intervals (Complicated N in range %). These values are shown for both the discovery and validation cohorts. We also show the prevalence of each adverse outcome and the associated confidence interval obtained via bootstrapping, for both validation and discovery cohorts. Threshold for statistical significance of the odds ratio was $p<9\times10^{-6}$ with Bonferroni correction.

| **Marker** | HCT | HGB | RBC | MCV |
|---|---|---|---|---|
| **Direction** | Increase | Increase | Increase | Decrease |
| **Threshold** | 0.261 | 0.667 | 0.071 | -0.747 |
| **Validation OR** | 1.45 [1.28, 1.64], p=3.192567e-09 | 2.03 [1.58, 2.6], p=2.0564184e-08 | 2.13 [1.75, 2.6], p=9.3e-14 | 1.51 [1.33, 1.71], p=6.6947e-11 |
| **Discovery OR** | 1.47 [1.24, 1.74], p=9e-06 | 2.28 [1.62, 3.2], p=2e-06 | 2.33 [1.77, 3.07], p=1.41e-09 | 1.41 [1.22, 1.63], p=3e-06 |
| **Discovery Total N** | 2153 | 328 | 526 | 3349 |
| **Validation Total N** | 4143 | 672 | 1033 | 3786 |
| **Discovery Complicated N** | 162 | 39 | 62 | 251 |
| **Validation Complicated N** | 327 | 76 | 118 | 332 |
| **Discovery Complicated N in range %** | 84 [78, 90] | 77 [64, 90] | 81 [71, 90] | 92 [89, 95] |
| **Validation Complicated N in range %** | 81 [77, 86] | 80 [70, 89] | 76 [68, 83] | 89 [86, 93] |
| **Discovery Prevalence** | 5.4 [5.2, 5.7] | 5.4 [5.2, 5.7] | 5.4 [5.2, 5.7] | 5.4 [5.2, 5.7] |
| **Validation Prevalence** | 5.7 [5.6, 5.9] | 5.7 [5.5, 5.9] | 5.7 [5.5, 5.9] | 5.7 [5.5, 5.9] |
| **Discovery PPV** | 7.5 [6.4, 8.7] | 11.9 [8.3, 15.3] | 11.8 [8.9, 14.5] | 7.5 [6.6, 8.3] |
| **Validation PPV** | 7.9 [7.1, 8.8] | 11.3 [8.8, 13.7] | 11.4 [9.5, 13.4] | 8.8 [7.8, 9.7] |
| **Discovery Recall** | 10.2 [8.7, 11.6] | 2.5 [1.7, 3.2] | 3.9 [2.9, 4.9] | 15.8 [14.1, 17.6] |
| **Validation Recall** | 11.3 [10.1, 12.5] | 2.6 [2.0, 3.2] | 4.1 [3.4, 4.8] | 11.4 [10.2, 12.6] |

| | | | | |
|---|---|---|---|---|
| **Discovery NPV** | 94.7 [94.5, 95.0] | 94.6 [94.4, 94.9] | 94.7 [94.4, 94.9] | 94.8 [94.6, 95.1] |
| **Validation NPV** | 94.5 [94.3, 94.7] | 94.3 [94.1, 94.5] | 94.4 [94.2, 94.6] | 94.5 [94.3, 94.7] |

**Table S18. Odds ratios and PPVs for rare behaviors are consistent in an out-of-sample validation cohort for preterm.** Out of sample validation of rare dynamics for hemoglobin HGB (g/dL), red blood cell count RBC ($10^6$/$mm^3$), hematocrit HCT (%), platelet count PLT ($10^3$/µL), and mean red cell volume MCV (fL) shows odds ratios, PPV, and NPV consistent with the discovery cohort. The table reports the total number of pregnancies with rare longitudinal change between 7-14 and 26-29 weeks' gestation (Total N), the total number of pregnancies with the complication of interest, the rare longitudinal change (Complicated N), and the percentage of pregnancies with both the complication and a marker value within the reference interval for 26-29 weeks' gestation with associated confidence intervals (Complicated N in range %). These values are shown for both the discovery and validation cohorts. We also show the prevalence of each adverse outcome and the associated confidence interval obtained via bootstrapping, for both validation and discovery cohorts. Threshold for statistical significance of the odds ratio was $p<9\times10^{-6}$ with Bonferroni correction.

| **Marker** | HCT | HGB | RBC |
|---|---|---|---|
| **Direction** | Increase | Increase | Increase |
| **Threshold** | 1.185 | 0.222 | 0.071 |
| **Validation OR** | 1.54 [1.33, 1.78], p=4.259079e-09 | 1.53 [1.32, 1.78], p=2.6255306e-08 | 1.76 [1.45, 2.13], p=6.308995e-09 |
| **Discovery OR** | 1.67 [1.35, 2.05], p=2e-06 | 1.64 [1.34, 2.01], p=2e-06 | 1.92 [1.47, 2.51], p=2e-06 |
| **Discovery Total N** | 986 | 1004 | 526 |
| **Validation Total N** | 2059 | 1889 | 1033 |
| **Discovery Complicated N** | 116 | 119 | 69 |
| **Validation Complicated N** | 237 | 221 | 135 |
| **Discovery Complicated N in range %** | 80 [73, 87] | 87 [81, 93] | 87 [78, 94] |
| **Validation Complicated N in range %** | 78 [73, 83] | 88 [83, 92] | 82 [76, 89] |
| **Discovery Prevalence** | 6.8 [6.5, 7.1] | 6.8 [6.5, 7.1] | 6.8 [6.5, 7.1] |
| **Validation Prevalence** | 7.4 [7.1, 7.6] | 7.4 [7.1, 7.6] | 7.4 [7.2, 7.6] |
| **Discovery PPV** | 11.8 [9.9, 13.9] | 11.9 [9.9, 14.0] | 13.1 [10.2, 16.2] |
| **Validation PPV** | 11.5 [10.2, 12.9] | 11.7 [10.3, 13.1] | 13.1 [11.1, 15.1] |
| **Discovery Recall** | 5.9 [4.8, 7.0] | 6.0 [5.0, 7.1] | 3.5 [2.7, 4.3] |
| **Validation Recall** | 6.4 [5.6, 7.2] | 5.9 [5.2, 6.7] | 3.6 [3.0, 4.2] |
| **Discovery NPV** | 93.4 [93.1, 93.7] | 93.4 [93.1, 93.7] | 93.3 [93.0, 93.6] |
| **Validation NPV** | 92.8 [92.6, 93.0] | 92.8 [92.5, 93.0] | 92.7 [92.5, 93.0] |
